# Colonoscopy polyp classification via enhanced scattering wavelet convolutional neural network

**DOI:** 10.1101/2024.04.17.24305891

**Authors:** Jun Tan, Jiamin Yuan, Xiaoyong Fu, Yilin Bai

**Affiliations:** School of Mathematics, Sun Yat-Sen University, Guangzhou, Guangdong, China; Guangdong Province Key Laboratory of Computational Science, Sun Yat-Sen University, Guangzhou, Guangdong, China; Health construction administration center, Guangdong Provincial Hospital of Chinese Medicine, Guangzhou, Guangdong, China; The Second Affiliated Hospital of Guangzhou University of Traditional Chinese Medicine(TCM), Guangzhou, Guangdong, China

## Abstract

Among the most common cancers, colorectal cancer (CRC) has a high death rate. The best way to screen for colorectal cancer (CRC) is with a colonoscopy, which has been shown to lower the risk of the disease. As a result, Computer-aided polyp classification technique is applied to identify colorectal cancer. But visually categorizing polyps is difficult since different polyps have different lighting conditions.

Different from previous works, this article presents Enhanced Scattering Wavelet Convolutional Neural Network (ESWCNN), a polyp classification technique that combines Convolutional Neural Network (CNN) and Scattering Wavelet Transform (SWT) to improve polyp classification performance. This method concatenates simultaneously learnable image filters and wavelet filters on each input channel. The scattering wavelet filters can extract common spectral features with various scales and orientations, while the learnable filters can capture image spatial features that wavelet filters may miss.

A network architecture for ESWCNN is designed based on these principles and trained and tested using colonoscopy datasets (two public datasets and one private dataset). An n-fold cross-validation experiment was conducted for three classes (adenoma, hyperplastic, serrated) achieving a classification accuracy of 96.4%, and 94.8% accuracy in two-class polyp classification (positive and negative). In the three-class classification, correct classification rates of 96.2% for adenomas, 98.71% for hyperplastic polyps, and 97.9% for serrated polyps were achieved. The proposed method in the two-class experiment reached an average sensitivity of 96.7% with 93.1% specificity.

Furthermore, we compare the performance of our model with the state-of-the-art general classification models and commonly used CNNs. Six end-to-end models based on CNNs were trained using 2 dataset of video sequences. The experimental results demonstrate that the proposed ESWCNN method can effectively classify polyps with higher accuracy and efficacy compared to the state-of-the-art CNN models. These findings can provide guidance for future research in polyp classification.

## Introduction

According to statistics [1] [2], colon and rectal cancers (CRC) are the most common types of cancers. Some polyps (adenomas) have the potential to develop into cancer. Therefore, it is crucial to detect and remove polyps from the body to mitigate the risk of cancer. Early diagnosis and removal of polyps significantly reduce the risk of CRC [3].

Colonoscopy is considered the gold standard for detecting and identifying polyps. The accuracy of classification depends on the skills and experience of the endoscopists. However, the diagnostic performance is limited, and up to 3.7% of CRC cases are post-colonoscopy or interval CRCs, which are CRCs diagnosed within three years after a normal colonoscopy [4]. One of the contributing factors to this issue is the prolonged duration of colonoscopy procedures, which can lead to mental and physical fatigue in human operators, resulting in degraded analysis and diagnosis. Other factors that may impact classification results include variations in illumination conditions, texture, appearance, and occlusion [5]. Additionally, since the appearances of different types of polyps are very similar, as depicted in Fig 1., distinguishing between various types of polyps can be challenging.

**Fig 1.**
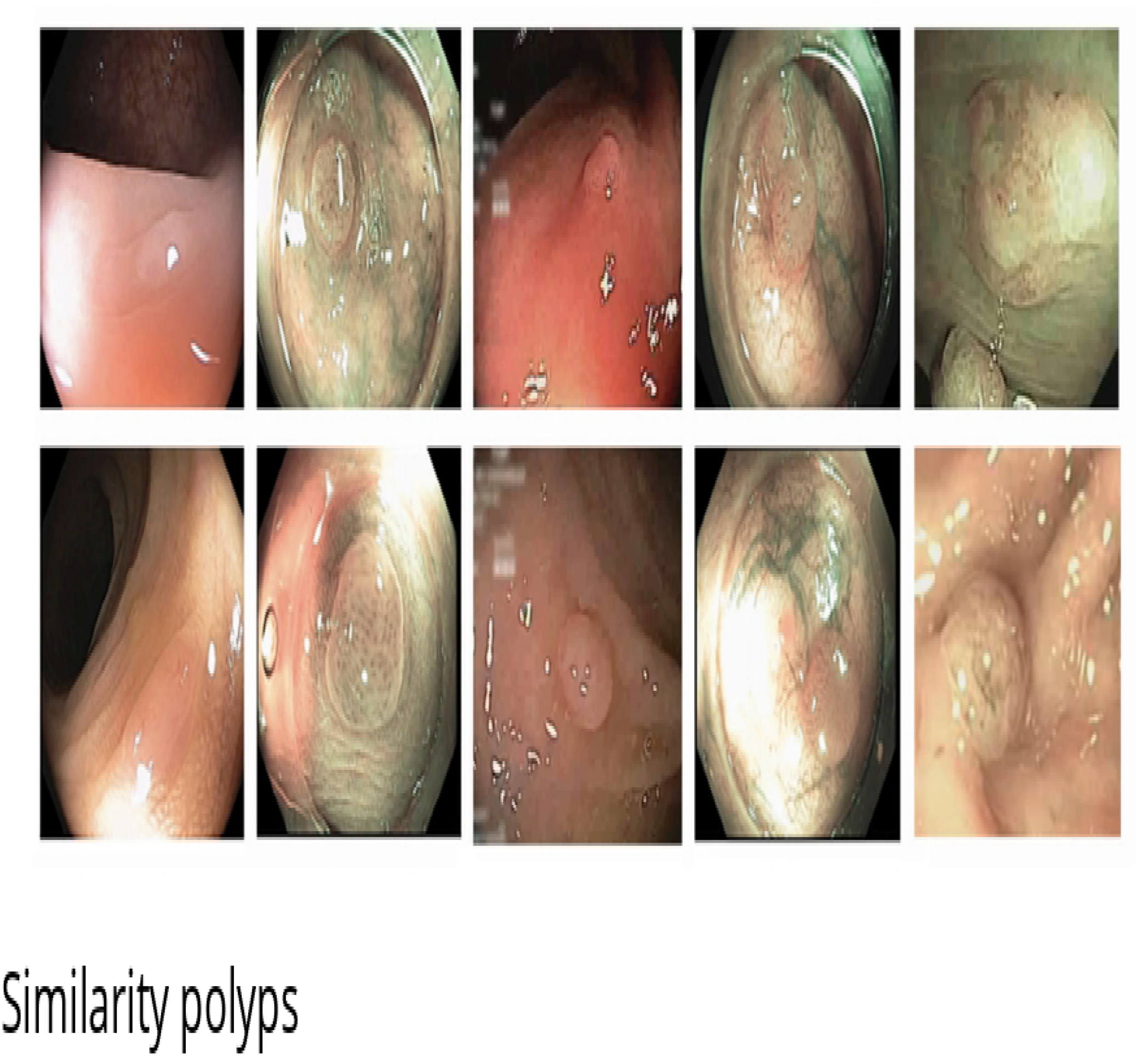
Similarity Upper: Five examples of adenomatous polyps. Lower: Five examples of hyperplastic polyps. The video images of these polyps are very simil ar, but they belong to differe11t classes (adenon1atous, hyperplastic).

In recent years, there has been a growing interest in the development of Computer-Aided Diagnosis (CAD) systems for automatic polyp detection and prediction of histology. The classification of polyp images has been achieved by CAD systems based on Machine Learning (ML). Tamaki et al. proposed a CAD system to classify colorectal tumors in Narrow Band Imaging (NBI) endoscopy using local features [6], achieving an accuracy of 96% on a 10-fold cross-validation using a dataset of 908 NBI images and 93% using an independent test dataset. It is important to note that these ML-based architectures consist of feature extraction and a classifier, and the systems require extensive preprocessing of the image datasets to extract the relevant features of the polyp images. Most ML-based methods have utilized Principal Component Analysis (PCA) [8] [9], Direction Discrete Wavelet Transform (DWT) [7], K-Nearest Neighbor (KNN) [29], and support vector machine (SVM) [12] based on handcrafted features. For example, the extraction of edge features detected in the images and their regions enables automated polyp detection via a classification system [11]. Local Fractal Dimension (LFD) [13] features extract shape and gradient information from the image to enhance the discriminativity of colonic polyps. In this present work, PCA and Wavelet are also used, and we will discuss these in the following section.

In contrast to ML-based methods that heavily rely on handcrafted feature extraction, Deep Learning (DL) has the advantage of not requiring previous preprocessing of image datasets, as they can be trained to automatically extract and learn the relevant features. As a result, a recent review compiled more than three hundred DL-based studies used in the field of medical image analysis [14], and related analysis revealed that the diagnostic performance of DL models is equivalent to that of healthcare professionals [15]. A significant amount of research on automatic polyp classification has been carried out since 2014. Remarkably, some participants in the Automatic Polyp Detection at the International Conference on Medical Image Computing and Computer Assisted Intervention (MICCAI) in 2015 [16] already used DL approaches. Since then, a growth of DL-based research accomplishing these tasks has been introduced every year with promising results [17].

Convolutional Neural Network (CNN) is a powerful technique in Deep Learning for medical image diagnosis. In contrast to traditional handcrafted feature extraction, CNN can effectively extract abstract and higher-level features [18]. Bernal et al. [16] compared the efficacy of handcrafted features with CNN-extracted features in detecting polyp presence on still frames. They claimed that end-to-end learning approaches based on CNN are more efficient than those based on handcrafted features. Akbari et al. [19] applied CNN on whole-slide images to classify informative and non-informative colonoscopy frames. Others have also utilized deep learning architectures, such as Visual Geometry Group (VGG) [20], for the identification of the existence of polyps. A comparative assessment of 11 CNN models has been performed for colorectal cancer two-stage classification. The CNN models include VGG16, VGG19 [20], Inception V3 [24], Xception, GoogLeNet [25], ResNet50, ResNet100 [26], DenseNet [27], NASNetMobile, MobilenetV2, for informative polyp frame detection [21]. Sharma et al. [22] proposed a multiple CNNs (ResNet, GoogleNet, Xception) classifier consultation strategy to create an effective and powerful classifier for polyp identification, achieving a performance measure greater than 95% in each of the algorithm parameters. Younas et al. [28] proposed an ensemble CNN-based approach for colorectal polyp classification, achieving a 96.3% F1 score on a public dataset.

Despite achieving good scores for polyp classification using CNN algorithms, one of the most significant limitations always present in the application of CNNs, especially in medical image analysis of colonoscopic videos and images, is the requirement for large, labeled datasets specific to the medical domain. Creating high-quality datasets in this domain is a challenge due to the high costs in terms of both economy, time, and medical expertise. In order to learn effective features for polyp classification, the depth and number of parameters in CNNs must be sufficiently large. However, due to the limited training samples for polyps, overly complex networks can easily lead to overfitting. Additionally, a complex network necessitates a lengthy training time. Other challenges in using CNNs for classification with limited sample data include working with smaller medical image datasets (at the level of thousands).

Recently, several methods have been proposed to tackle the issue of limited training samples in deep learning-based image classification. Given that wavelets can extract effective features from images even with small sample sizes, some approaches combining wavelet and CNN for image classification tasks have been introduced.

Razali et al. [30] integrated wavelet with CNN for breast tissue classification to address the problem of CNN overfitting. Simon et al. [31] introduced an architecture called WaveTexNeT, which combines wavelet and the Xception convolutional network. This model concatenates spatial and spectral features as inputs for the network. However, WaveTexNeT utilizes spectral features only as a data augmentation technique, instead of using original pixels as network inputs. Deo et al. [32] developed an ensemble model incorporating wavelet and CNNs. They extract features from images using 2D empirical wavelet transform, with CNNs employed for image classification. Nevertheless, the ensemble model still contains a considerable number of learnable parameters. Kutlu et al. [33] devised a novel method based on CNN, DWT, and SVM for polyp detection and classification. In this approach, DWT is utilized to reduce the dimensionality of feature vectors obtained from CNNs. However, the wavelet method only learns the spectral coefficients of CNN features, including approximations and details, which limits the full exploitation of backpropagation to optimize the entire CNN network.

In this article, we introduce a novel network named Enhanced Scattering Wavelet Convolutional Neural Network (ESWCNN) to effectively integrate wavelet transform with the standard convolutional network. Unlike existing methods that primarily utilize Discrete Wavelet Transform (DWT) for preprocessing [32] or postprocessing [33], and those neglecting to train the standard convolutional network’s learnable parameters, our proposed approach processes each input channel through a fixed wavelet layer alongside the original image layers. This is followed by the application of learnable 1 *×* 1 filters to generate the output channels. Building upon ESWCNN, we have devised an architecture for polyp classification that not only captures deep spatial features but also extracts spectral information. This architecture is an end-to-end model, eliminating the need for an additional classifier.

The key contributions of this article are outlined as follows:

1) In order to extract more discriminative features and develop an end-to-end system efficiently with a limited number of polyp samples, we propose a new method called Ensemble ML and DL. This method enables the network to extract deep features effectively with fewer trainable parameters that can be learned from a small training dataset. The scattering wavelet is capable of extracting common spectral features with various scales and orientations, while the CNN learnable filters can capture spatial features that DWT may overlook.

2) We suggest incorporating local discriminant structure into the cross loss function by combining the scattering wavelet and CNN losses. This approach aims to enhance the learning of more discriminative features and establish an end-to-end system simultaneously.

3) Our proposed method outperforms other state-of-the-art polyp classification techniques significantly, especially when dealing with limited training samples. Furthermore, due to its simple structure, our method exhibits faster training and testing speeds compared to the current state-of-the-art methods.

## Related works

Numerous researchers have conducted studies on the diagnosis of polyps from various perspectives. However, there is ample room for enhancing diagnostic performance. Previous research on polyp diagnosis can be broadly categorized into three main areas: detection, segmentation, and classification. In comparison to other domains, the classification of polyps has received less scrutiny. Polyp classification studies have utilized various technologies, including computer vision, machine learning, and deep learning.

Based on the results of previous research and the findings of the MICCAI Endoscopic Vision Challenge [16], it is evident that state-of-the-art object detection models can already achieve very high precision in polyp detection. In this paper, we assume that the polyps have been detected and narrow our focus to the study of classification.

Several models have been proposed for the automated classification of colon polyps. Mesejo et al. [43] suggested a model that combines machine learning and computer vision algorithms to perform a virtual biopsy of hyperplastic lesions, serrated adenomas, and adenomas. They also introduced a dataset of colonoscopic videos with ground truth collected from experts, referred to as the colonoscopy dataset, which includes 76 videos presented in both White Light (WL) and Narrow-Band Imaging (NBI) formats. The NBI video format was utilized in the study, with the dataset containing 15 serrated, 21 hyperplastic, and 40 adenoma polyps. These videos comprise 20,948 adenoma, 7,423 hyperplastic, and 5,902 serrated polyp images, evaluated by four experts and three beginner operators. The study combines the advantages of both computer vision and machine learning to achieve accurate classification. However, the average accuracy (ACC) achieved is 82.46%, with a sensitivity (SEN) of 72.74% and a specificity(SPE) of 85.88%. The experiment compares the 15 best-ranked models, with the top-performing model utilizing Random Subspaces (RS) or Support Vector Machine (SVM) considering WL, 3D shape, color, and textural features.

Wavelets have wide applications in signal processing, pattern recognition, and other fields due to their superior performance in time-frequency analysis. Some Wavelets include the following:

1) Db97 [34]: The Db series wavelet is a family of wavelets proposed by Donoho and Johnstone, also known as “Daubechies wavelets”. Db 97 refers to a wavelet of order 9, with a filter length of 97 coefficients. Db wavelets exhibit good orthogonality and symmetry, making them popular choices in applications such as signal denoising and image compression.

2) Bior39 [35]: The Bior series wavelets combine the features of orthogonal and biorthogonal wavelets. Bior39 is constructed using three db wavelets (db 2, db 4, db 6) and three symmetric wavelets (sym 2, sym 4, sym 6). Bio wavelets are frequently employed in biomedical signal processing and image compression applications.

3) Sym5 [36]: The Sym series wavelet is a type of biorthogonal wavelet family. Sym 5 is created using five db wavelets (db 2, db 4, db 6, db 8, db 10). The Sym wavelet exhibits good approximate symmetry and is well-suited for signal processing and image compression tasks.

4) Db4 [34]: Db 4 belongs to the Db series of wavelets with a filter length of 4. The Db4 wavelet is a popular choice among discrete wavelets, extensively employed in tasks such as signal denoising and image compression, thanks to its short filter length and excellent time-frequency localization properties.

HHT [37]: Hilbert-Huang transform(HHT), consisting of empirical mode decomposition and Hilbert spectral analysis, is a newly developed adaptive data analysis method, which has been used extensively in signal processing. The HHT transform is usually used for image processing, especially in image compression, which is able to provide a better compression effect and a faster computing speed.

For image classification tasks, CNNs are susceptible to noise interference. To address this issue, several methods have been developed that integrate CNNs with wavelets. One such method is the Multi-level Wavelet CNN (MWCNN) [53], which incorporates wavelet transform into the CNN architecture to reduce the resolution of feature maps. Another approach is WaveCNet [54], which integrates CNNs with wavelets by replacing the conventional pooling layer with discrete wavelet transform. This allows the wavelet to decompose the feature maps into low-frequency and high-frequency components. By integrating wavelets with commonly used CNNs such as ResNet [26], DenseNet [27], and VGG [25], higher accuracy in image classification tasks has been achieved. One drawback of the aforementioned methods is that they directly replace the pooling layer with wavelets, leading to the loss of spatial feature information.

In WaveTexNeT [31], pooling and the convolution operation are considered as downsampling. The frequency domain provides an advantage for feature extraction. By enhancing specific frequencies and suppressing others, a spatial filter can be easily made selective. In CNNs, controlling this selection is challenging. WaveTexNeT incorporates spectral techniques into CNNs to extract spectral and spatial features, but the deep learning network only utilizes Xception.

The CNN-Wavelet scattering textural feature fusion method [30] is similar. It aims to address CNN overfitting on small datasets by incorporating scattered wavelet coefficients to preserve high-frequency signal information. The classifier in this approach employs a non-parametric KNN algorithm. Therefore, CNN-Wavelet fusion is not an end-to-end solution.

In the Colonoscopy Dataset [43], Kutlu et al. [33] introduced a novel approach for polyp detection and classification using CNN, DWT, and SVM. The method involves ensemble CNNs for feature extraction, DWT for feature reduction, and SVM for polyp classification. The experiments were conducted using 5-fold cross-validation. The study not only classified the three basic classes of polyps - Serrated adenomas, adenomatous polyps, and hyperplastic polyps but also introduced a lumen class to reduce incorrect estimates in polyp detection.

## Materials and methods

To implement the proposed method ESWCNN in this study, the experiment is conducted in three stages, Fig 2 illustrates the schematic diagram of the feature extraction method using ESWCNN. Firstly, the entire input images of the polyps undergo pre-processing. Texture images can be processed for both spatial and spectral features. The motivation behind the ESWCNN method stems from the limitation of CNNs in capturing spectral information essential for processing texture images. Spatial features provide detailed information extracted from textures by manipulating pixel intensity values in the neighborhood. DWT [7] has shown promise in capturing spectral features, encompassing approximation features like low and high frequencies. The low frequency component depicts the smooth areas, while the high frequency component captures spectral features like edges and boundaries [31]. Therefore, spatial-spectral features from texture images are combined and fed into the network for training.

**Fig 2.**
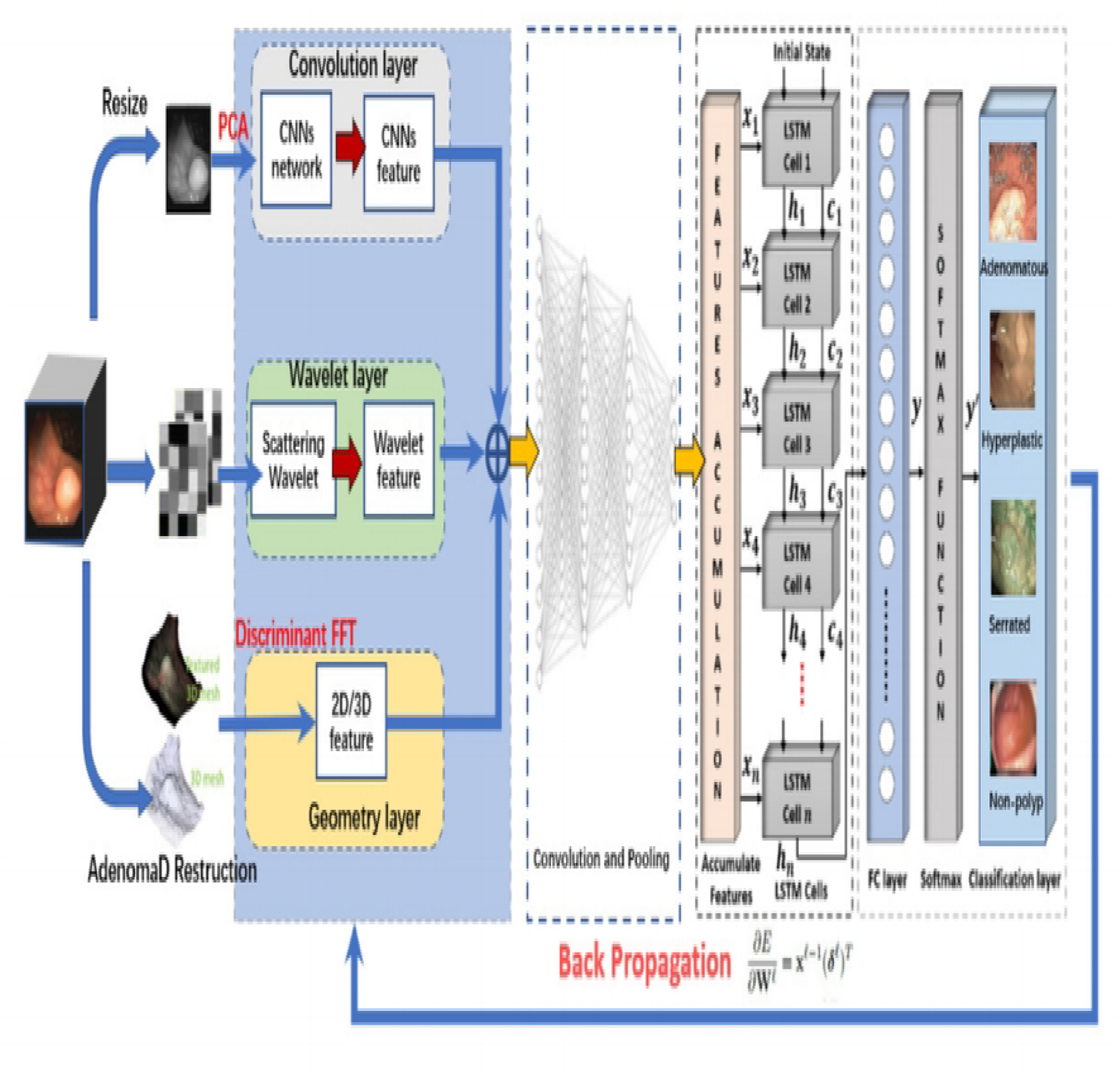
Proposed architecture for ESWC N for polys classification.

Secondly, PCA [9] is utilized to reduce the dimensionality of the spatial-spectral feature vector space. The subsequent section will elaborate on how PCA can enhance the performance of texture classification.

Finally, CNN is employed as a parallel classification algorithm on the input feature patches extracted from the previous stage. The accuracy of the proposed classifier models is evaluated using confusion matrices, precision, recall, and classification accuracy.

### 2D/3D Feature Extraction

For the 2D texture feature extraction, only a single region of interest from a frame where the lesion is visible is required. This region of interest does not need to be highly precise and can be manually defined as a simple polygonal region. Invariant Local Binary Patterns (ILBP) [55] and Invariant Gabor Texture Descriptors (AHT) [56] are chosen as texture descriptors for this purpose. These descriptors are selected for their robustness against monotonic gray-scale changes, such as those caused by variations in illumination, and for their rotational invariance. Gray-level co-occurrence matrix (GLCM) or Histograms of Oriented Gradients (HOG) descriptors are not utilized because these features, in their standard form, are not invariant to rotation or scale changes in the texture. In endoscopy, the light source is typically positioned very close to the camera’s center of projection. Therefore, a pixel classified as a specularity indicates that the normal at that point of the surface aligns with the optical beam. Consequently, lesions with irregular shapes exhibit distinct specular patterns.

We rely on Agisoft Metashap software Structure-from-Motion(SfM) [48] to compute a dense 3D model of the polyp and the surrounding tissue (see Fig 3) from the exploratory video.

**Fig 3.**
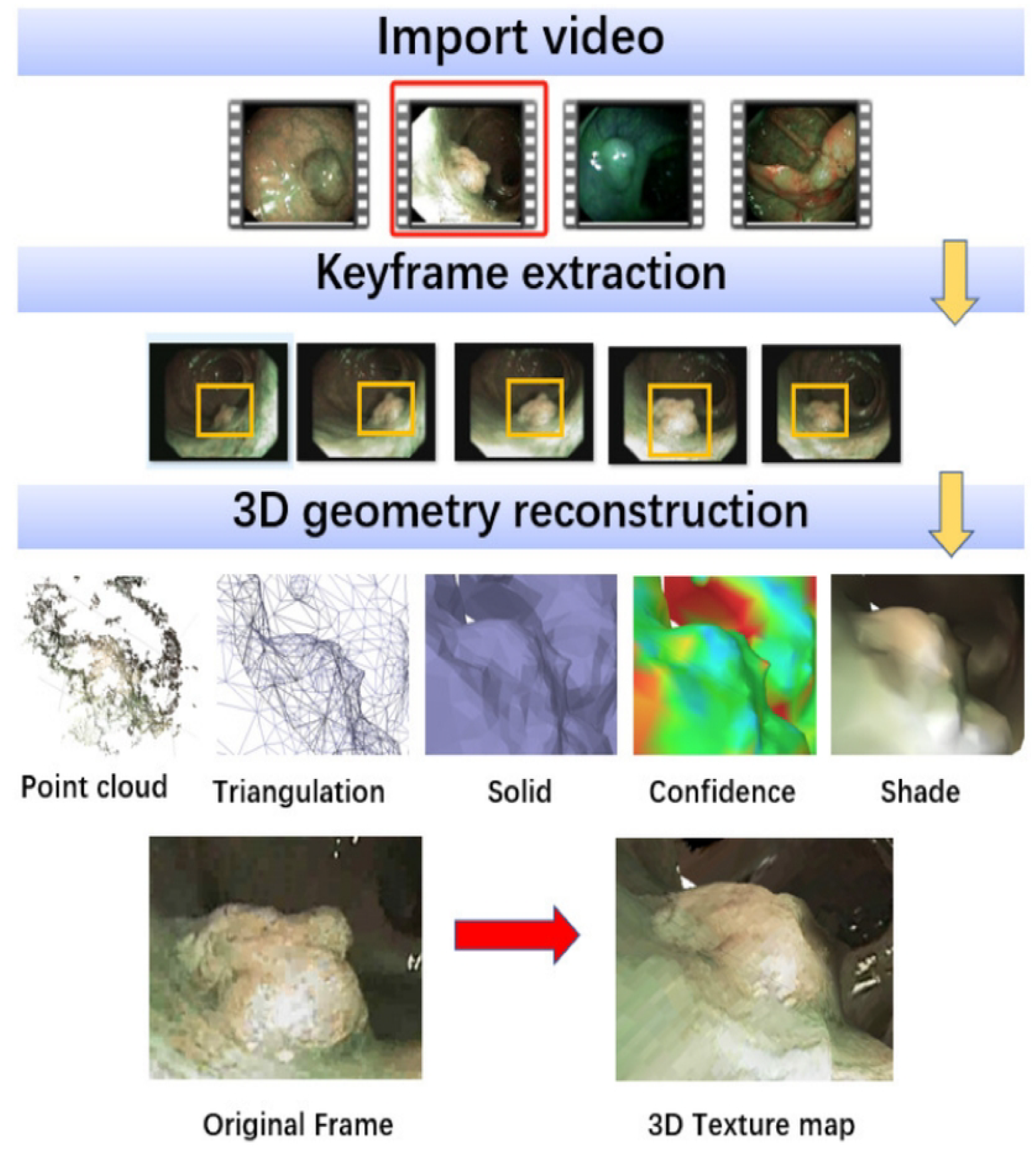
3D reconstruction using SflvI.

To achieve accurate and stable reconstructions, current Structure-from-Motion (SfM) methods typically require the following conditions:

1. Rigid Geometry: While natural deformations may occur in colon tissues (e.g., due to peristalsis or external compression), it is assumed that during the exploratory video, deformations near the target polyp are minimal, and the rigidity assumption holds true.
2. Textured Surfaces: Colon tissue, in general, lacks strong texture, which can impact the quality of 3D reconstruction. However, with Narrow Band Imaging (NBI) lighting, near-surface vessel patterns are highlighted, improving the textural content for reconstruction.

In the context of the SfM process described, the researchers utilize PhotoScan software [48], which automatically generates a dense, textured 3D mesh of the polyp from a set of images. This dense SfM reconstruction provides a detailed description of the polyp’s 3D surface using a triangular mesh(see Fig 3). This enables the computation of the geometric quantities such as normals or curvatures. The features extracted from the reconstructed 3D surface, along with their corresponding dimensionalities, are summarized in Table 1. This table likely provides a comprehensive overview of the key characteristics and properties extracted from the reconstructed polyp surface.

**Table 1.** Summary all feature descriptors and their corresponding size of feature vector space [43].

| Feature | Descriptor | Number of features |
| --- | --- | --- |
| 2D Texture | AHT(Invariant Gabor Texture) | 166 |
|  | Rotational Invariant LBP | 256 |
| 2D Color | Color Naming | 16 |
|  | Discriminative Color | 13 |
|  | Hue | 7 |
|  | Opponent | 7 |
|  | color GLCM | 33 |
| 3D Shape | Shape-DNA | 100 |
|  | Kernel-PCA | 100 |

Another function of the SfM software is to extract single-frame images from the video, a process that is elaborated on in the experimental section.

### Invariant Scattering Wavelets

A wavelet transform commutes with translations, and therefore is not translation invariant. The Discrete Wavelet Transform (DWT) [7] decomposes data into various components, separating the main information and details. The original data can be reconstructed using Inverse DWT (IDWT) with the DWT output. In signal processing, DWT is a valuable tool for anti-aliasing. In this paper, we primarily focus on its application in enhancing the spectral effect in Convolutional Neural Networks (CNNs) for polys image classification.

In the early studies of wavelet integrated neural networks, researchers implemented wavelet transforms using parameterized one-layer networks and searched for the optimal wavelet in the parameter domain. Recent work [32] has extended this method to deeper networks for image classification. However, training a deep network with wavelet parameterization is challenging due to the significantly increased computational complexity [32].

Mallat et al. explored the optimal deep network from a mathematical and algorithmic perspective, they introduced Scattering Wavelets (ScatNet) [44] by cascading wavelet transform with average-pooling and nonlinear modulus operation. Invariant Scattering Wavelets preserve image detail information and extract a translation invariant feature robust to deformations. Compared with CNNs of the same period, ScatNet achieves better performance on texture discrimination and recognition tasks.

Additional translation invariant coefficients *U* can be computed by further iterating on the Scattering wavelet transform, where modulus operators are defined as follows:

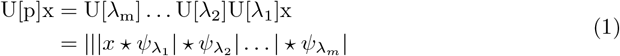

where index *λ* is the frequency location of *ψ*_*λ*_, and defines a path as squence *p* = (*λ*_1_, *λ*_2_, ;*λ*_*m*_) . To obtain scattering coefficients *S*, It defines a windowed scattering transform use a low pass filter 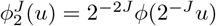

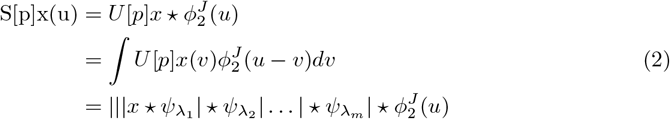

However, ScatNet is essentially a hand-designed feature extractor without learnable parameters. Due to the strict mathematical terms, ScatNet can not be easily transferred to image-to-image tasks, such as image segmentation.

To overcome this, the Wavelet feature is computed by applying a series of wavelet transforms to the image, and then averaging the results over a range of scales and orientations through the iterative and interconnecting process of averaging and wavelet filtering. Fig 4 shows the process of Scattering Wavelets from n number of levels to get the final feature through the combinations of *S* coefficients.

**Fig 4.**
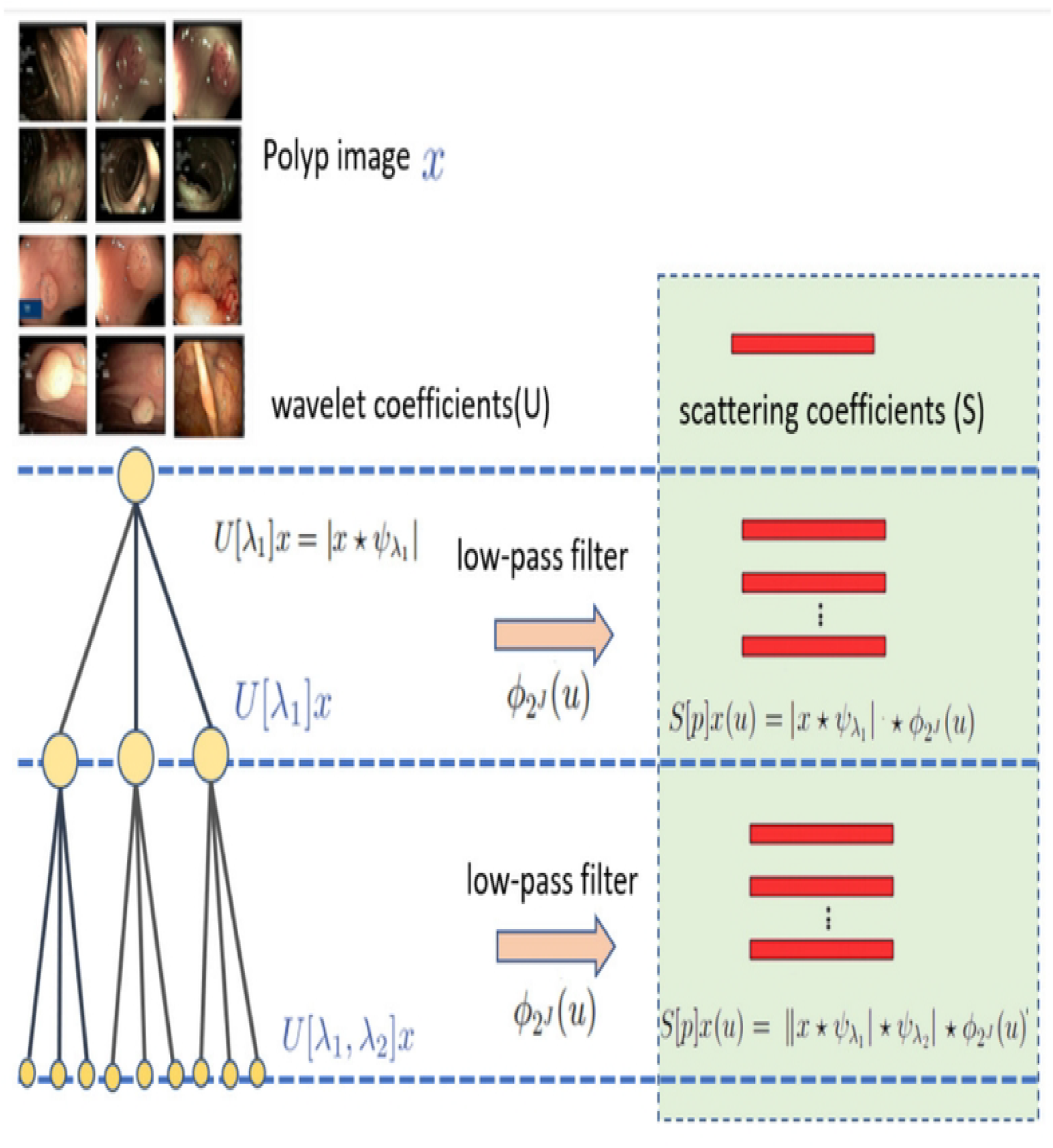
Wavelet scattering coefficients (S) i11 three WS levels for the feature extracti process.

ESWCNN is a encoder-decoder model implementing wavelet package transform (WPT) for image classification and process the concatenation of various components of the input data in a unified way. In ESWCNN, the input images are represented as four multiresolution levels of decomposition to extract better texture features. scattering wavelet extract features in multiresolution analysis in frequency domain. ESWCNN uses the 3 *×* 3 convolutional kernels, stride 2 with padding 1 *×* 1 for capturing the spectral features. Stride and padding is applied to the input image to lower the feature dimensions.

In ESWCNN, pooling and the convolution operation are considered as down sampling and filtering thereby establishing a relation between convolutional neural networks and multiresolution decomposition. The Invariant scattering frequency domain offers an advantage for feature extraction. By increasing certain frequencies while suppressing others, a spatial filter may be readily made selective. In CNNs, this explicit selection of specific frequencies is difficult to regulate. ESWCNN incorporate spectral techniques into CNNs through multiresolution analysis.

### Adaptive Principle Component Analysis

Principal Component Analysis (PCA) [10] is a mathematical technique for data transformation that reduces multidimensional data into a lower number of principal components, which are uncorrelated and retain variance as much as possible. PCA is often used for feature selection to address the issue of dealing with numerous features. This analysis reduces the feature dimension while minimizing information loss.

In the field of medical imaging, PCA has been employed for various purposes. For instance, Ansari et al. [39] used PCA to transform endoscopic narrow-band images (NBI) into standard colored endoscopic images, allowing for the extraction of a target image from a different source image.

In the context of extracting 3D shape features, PCA plays a crucial role in capturing fundamental information in NBI, which enhances structures and textures. This enables the improvement of standard images for better assessment and diagnosis. Additionally, PCA is used for reducing highly dimensional data, such as fluorescence spectral images of colorectal polyps. By reducing a high-dimensional set of images to eight principal components, tissue classification becomes more intuitive and manageable.

### Discriminant FFT-filter

The Fast Fourier Transform (FFT) [41] is an efficient algorithm for calculating the discrete Fourier transform (DFT). It can convert a signal from the time domain to the frequency domain and vice versa. While FFT is distinct from the wavelet transform, it holds significant importance in signal processing and is frequently used in conjunction with the wavelet transform to enhance the efficiency and effectiveness of signal processing.

CNN operators primarily focus on feature aggregation rather than adjusting specific frequency components. To address this limitation, we propose the integration of a discriminant operator that can effectively filter out various components. This integration involves incorporating a Discriminant Fast Fourier Transform filter (Discriminant FFT-filter) into the CNN network to extract useful maps while suppressing noise components in the frequency domain.

Specifically, given the encoder feature *F ∈ R*^*H×W ×C*^, we initially apply a 2D FFT*ℱ* operation along the spatial dimensions, resulting in a transformed feature *F*_*c*_ = *ℱ*[*F*] where *F*_*c*_ *∈ C*^*H×W ×C*^ . Subsequently, to learn a discriminant spectrum filter, we introduce a learnable weight map *W ∈ C*^*H×W ×C*^ and perform element-wise multiplication of *W* with *F*_*c*_. This spectrum filter enhances the training process by enabling global adjustments to specific frequencies, with the learned weights tailored to discriminate between different frequency components of the target distributions.

The output feature *F*_*out*_ is defined as:

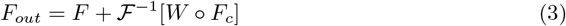

where *°* represents the hadamard product.

### Long Short Term Memory

In the second stage, Long Short Term Memory(LSTM), a variant of the RNN model [42], was used to exploit the temporal information from the set of *t* features vectors that were extracted in the first stage by using ResNet18. LSTM with optimized network parametrers was used to classify colorectal polyp [46].

In this work, we apply LSTM to analysis the features of signal, the signal regard as a time squence. The basic structure of a standard LSTM cell is shown in Fig 5, which illustrates the flow of data at time **t**. In general, four components, named as input gate (**i**_**t**_), forget gate (**f**_**t**_), cell candidate(**g**_**t**_), and output gate (**o**_**t**_), are responsible for controlling the state information at time step **t**.

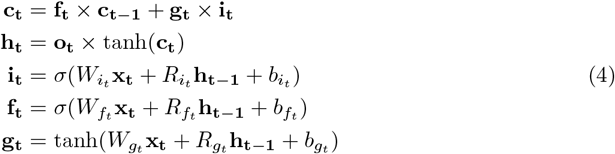

where tanh is the hyperbolic tangent function, and *σ* is the sigmoid function, which is used to compute the activation function of the gate. The (**i**_**t**_) controls the level of the cell state update, whereas the gate (**f**_**t**_) controls the level of the cell state reset. The (**g**_**t**_) adds the information to the cell state and finally, the (**o**_**t**_) controls the level of the cell state added to the hidden state. Based on these components, the complete structure of the cell is divided into three gates, named as forget, input, and output gates, as highlighted in Fig 5.

**Fig 5.**
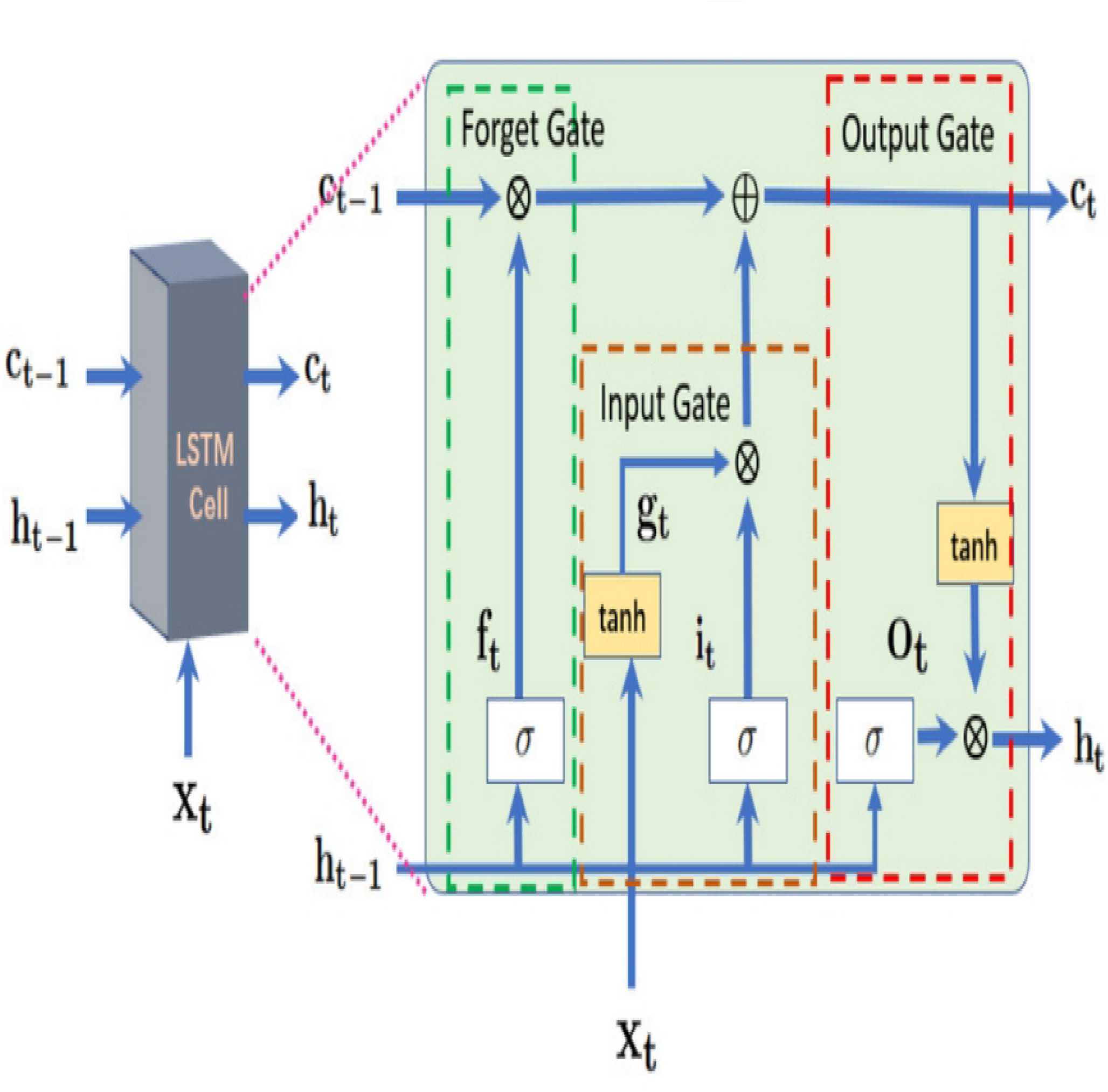
Internal connectivity of a standard LSTM cell -

### Scattering Convolutional Neural Network

Our proposed classification framework consists of a cascaded CNN and wavelet-based deep networks capable of classifying the video data based on spatiotemporal features. The primary advantage of our network is its capability to categorize a variable length sequence of n successive images (i.e.,*x*_1_, *x*_2_, *x*_3_, *x*_*j*_,;*j ∈ Z*) with significant performance gain. the l -th layer features 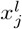 are obtained by l *−* 1 -th layer features 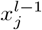.

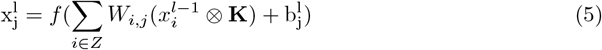

where **K** is convolutional kernel, ⊗ denotes convolution operation, activation function *f* computed hyperparameter *W*_*i,j*_ with bias *b*. For example, the use of more successive images results in better classification performance.

Furthermore, our cascaded deep learning model has exhibited superior performance compared to models solely based on CNNs. This can be attributed to the fact that CNN models typically focus on extracting spatial information by analyzing each input image separately, without taking into account both spatial and temporal features when dealing with video datasets. Due to the absence of temporal information consideration in CNN models, there is a degradation in the overall classification performance.

To overcome the limitation of previous spatial features-based methods in the medical domain, our study included a spatial variant of scattering wavelet*S*[*x*] along with the conventional CNN model to enhance the classification performance.

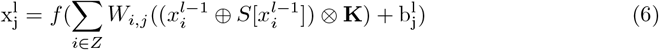

where ⊗ denotes that all groups are concatenated along the depth dimension. To remove the negative influence of autoencoder, we skip the autoencoder and updated parameter *W*_*i,j*_ by the gradient:

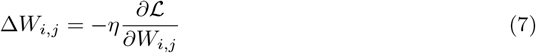

where *η* is learning rate, and loss function ℒ can be formulated as

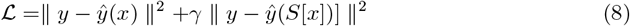

where *y* is ground truth class label, and ŷ(*x*) is predicted label, ŷ (*S*[*x*]) is a predicted label generated by scattering wavelet. Additionally, *γ* is a predefined weight used to balance the losses from wavelet scattering and CNN. Through experiments, we will demonstrate that selecting a suitable value for *γ* to achieve optimal performance is a straightforward task. Specifically, When*γ* = 0 or*γ→* 1 the performance tends to deteriorate. Further details regarding this observation will be discussed in the subsequent experimental section.

## Experiment

### Data set preparation

In this study, we have gathered all publicly available endoscopic datasets within the research community, in addition to curating a new dataset sourced from the University of Kansas [45]. All datasets have been deidentified to ensure patient confidentiality. Collaborating with endoscopists, the polyp classes were meticulously annotated across all collected video sequences, along with delineating the bounding boxes of polyps in each frame. The following provides an overview of each dataset.

The PolypGen dataset is a comprehensive collection designed for polyp segmentation and detection generalization [47]. Some representative negative and positive sample images are illustrated in Fig 6. This dataset comprises a total of 8037 frames, encompassing both individual frames and sequences. It includes 3762 positive sample frames and 4275 negative sample frames sourced from six distinct hospitals, each with diverse population demographics, endoscopic systems, surveillance expertise, and polyp resection techniques.

**Fig 6.**
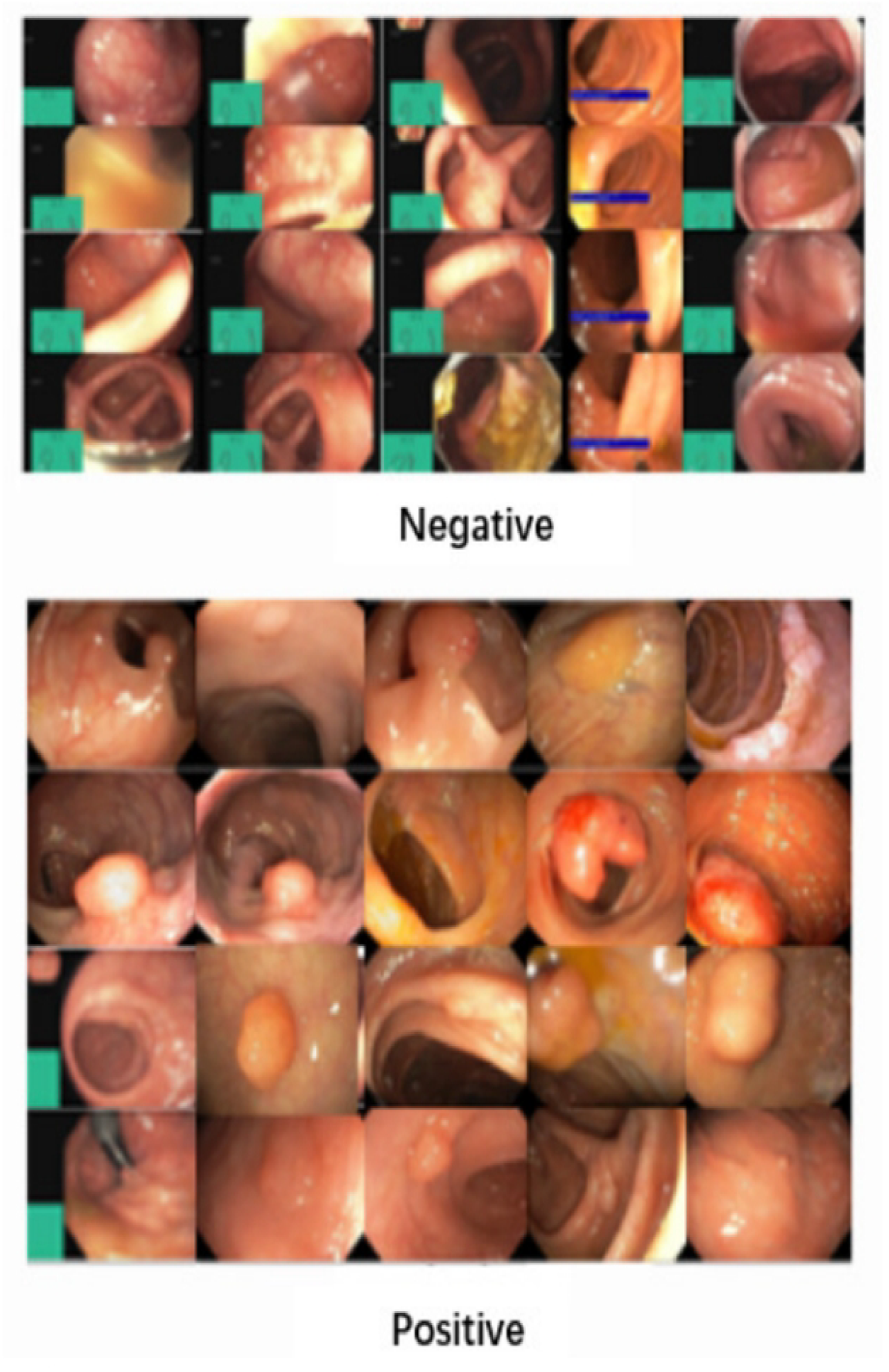
The polyps’ samples negative and positive classes of Pol_yp_Gen dataset

A portion of this dataset was initially employed in the 3rd International Workshop and Challenge on Endoscopic Computer Vision. This challenge aims to foster collaboration, curate multicenter datasets, facilitate the development of generalizable models, and evaluate deep learning techniques. The dataset provided represents an expanded iteration of the EndoCV2021 challenge.

The GLRC UCI dataset [43] is a publicly available dataset that focuses on Gastrointestinal Lesions in Regular Colonoscopy. This dataset comprises 76 short video sequences with class labels. Some sample images are illustrated in Fig 7 . The training and test data for the UCI model were derived from this dataset and classified by experts.

**Fig 7.**
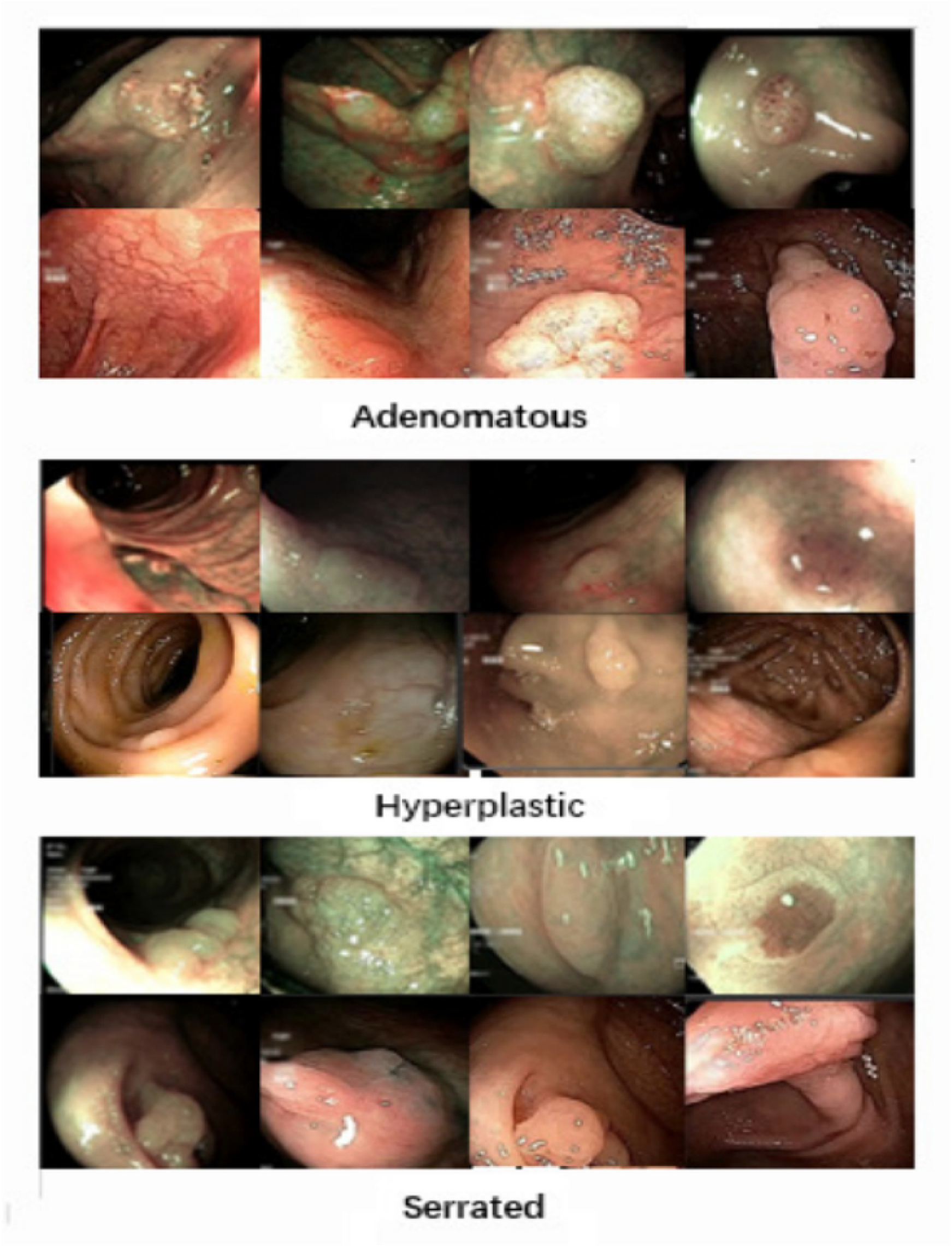
The polyps’ samples from different classes of UCI dataset

The dataset includes video sequences captured in both White Light (WL) and Narrow-Band Imaging (NBI) formats. It consists of 3 classes: 15 serrated, 21 hyperplastic, and 39 adenoma polyp videos. These videos contain a total of 20,948 adenoma frame images, 7,423 hyperplastic frame images, and 5,902 serrated polyp frame images, each captured from various angles.

For the image classification task, a subset of 1,200 images was selected, with 400 images from each class captured from different perspectives.

The GDZY dataset was gathered from the Second Affiliated Hospital of Guangzhou University of Traditional Chinese Medicine(TCM). It comprises 1,347 patient colonoscopy sequences. Due to the correlation between intestinal polyps and human magnetic signals [57], data were collected using a human weak magnetic signal instrument without gastroenteroscopic intervention.

Employing a paired design, the same subjects underwent colonoscopy both before and after colorectal polyp resection, with the paired images displayed in Fig 8. Following the paired results, Gastroenteroscopy experts manually labeled the polyp classes (negative or positive) for the entire dataset.

**Fig 8.**
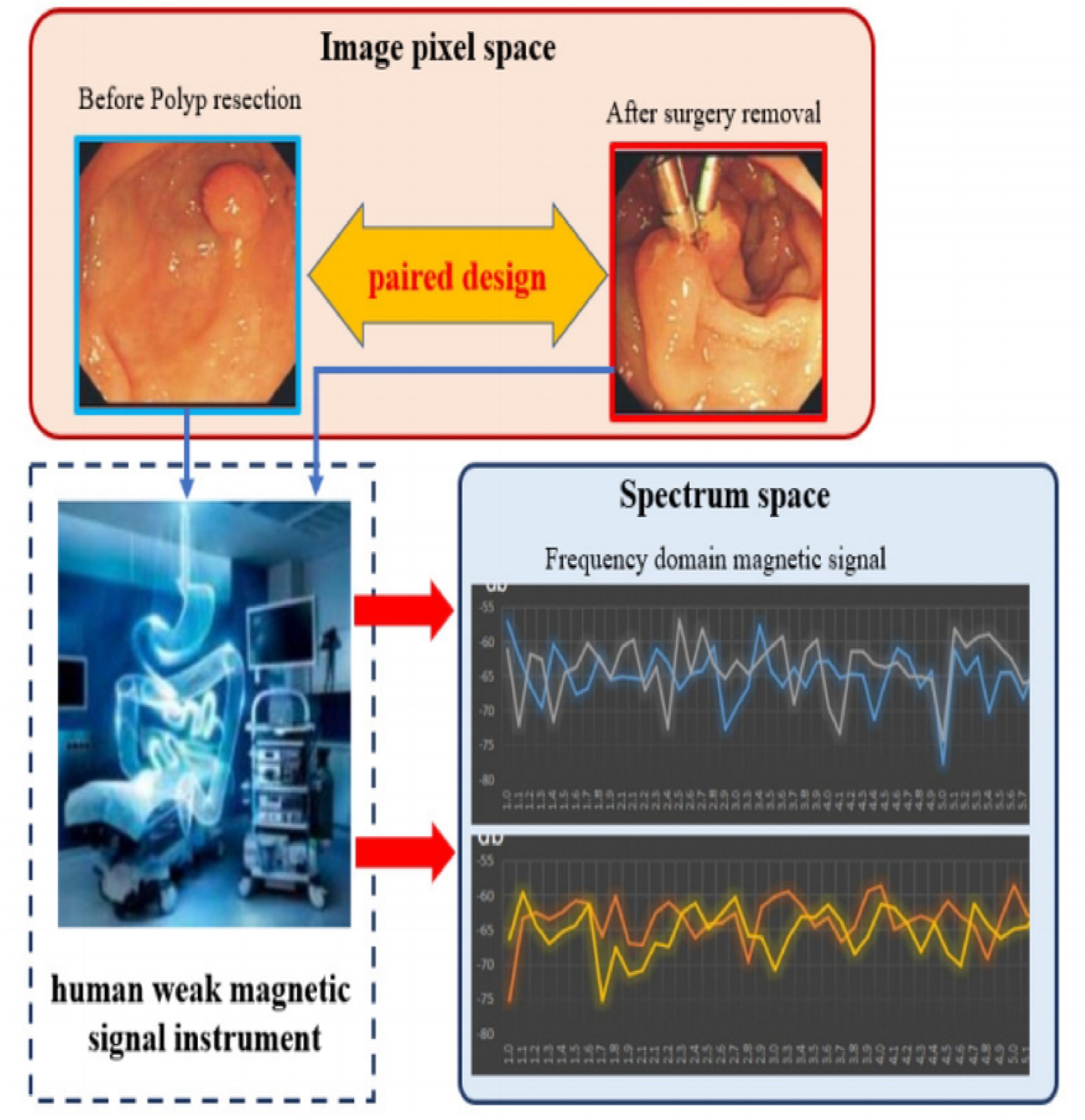
The polyps’ paired design of GDZY dataset.

In signal processing, wavelet transform is a method for analyzing the time-frequency characteristics of signals. After removing columns with zero values and columns of the same level, 715 electromagnetic wave variables were obtained. Fourier wavelet transformation was performed, including db 97 wavelet [34], HHT [37], Bior39 [35], Sym [36], db 4 [34], FFT [41], resulting in a total of 4,625 feature columns.

The following methods were used for feature selection:

1) Select the top 30 features based on feature importance from random forest. 2) Choose the top 99% features based on absolute correlation with the target variable. 3) Select the top 99% features based on the variance after softmax with the target variable. Combine the features selected by these methods through voting to obtain the common variables as the result of feature selection, which will be used as the overall features for the next algorithm construction.

The top 1 % significant features were identified by conducting a two-sample T-test on different features. In two groups of data divided by whether or not the individuals have colorectal polyps, if a feature significantly influences the presence of colorectal polyps, then this feature should exhibit a significant difference between the two groups (with and without colorectal polyps). Therefore, a two-sample T-test was used to assess whether the mean of the feature is significantly different, filtering out variables with high significance for further analysis.

Selected top 1% features shown in Fig 9 .

**Fig 9.**
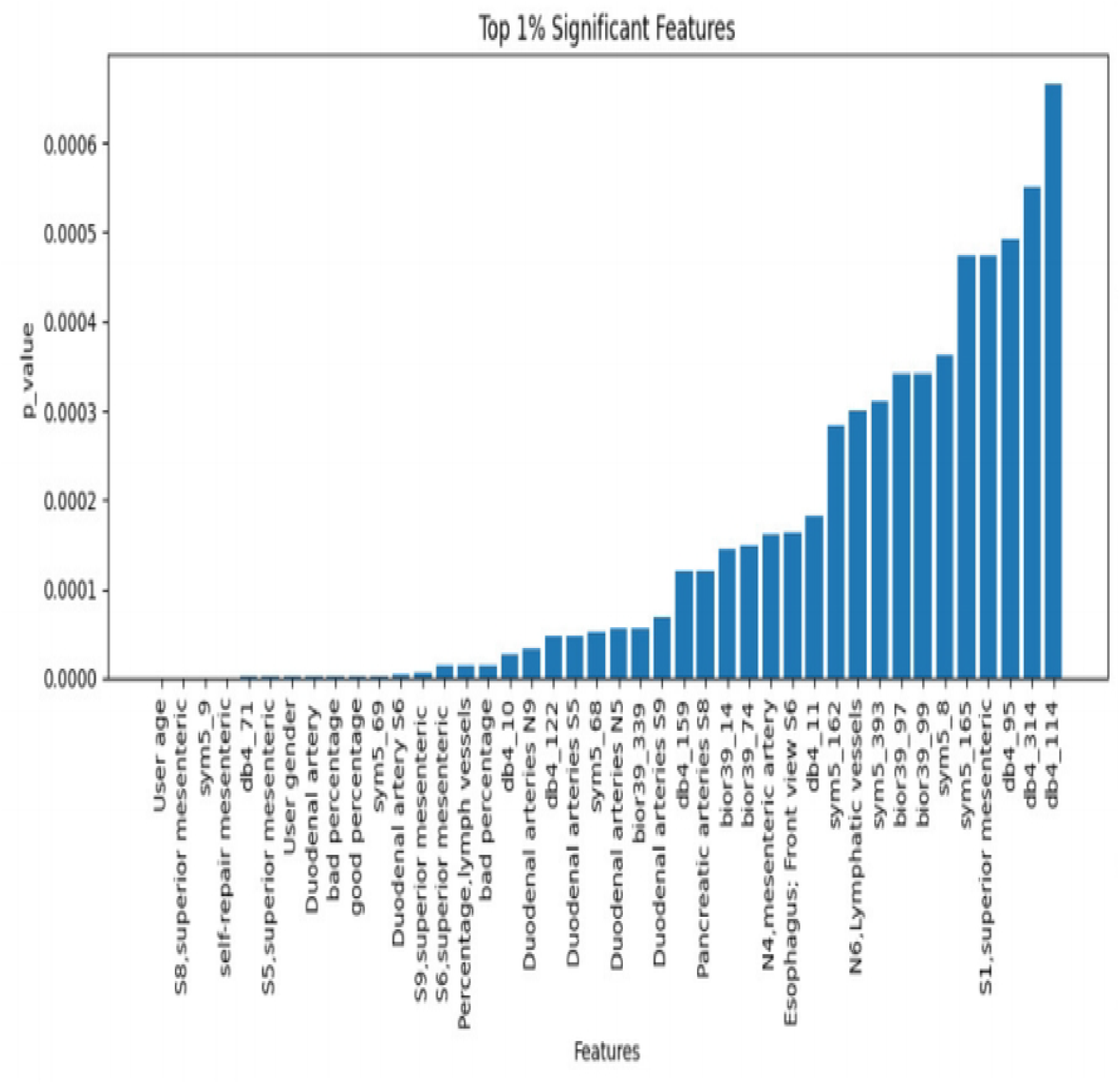
Top 1 significant features of GDZY dataset.

### Implementation Details

The proposed framework was implemented with MATLAB R2021a (MathWorks, Inc., Natick, MA, USA) on a Windows 11 operating system. The deep learning library named as deep learning toolbox was included in MATLAB for the implementation of various CNN models. All the experiments were performed on a desktop computer with a 3.50 GHz Intel (Santa Clara, CA, USA) Core-i7-10700K central processing unit (CPU), 32 GB random access memory (RAM), and an NVIDIA (Santa Clara, CA, USA) GeForce RTX 2060 graphics card . The graphics card was utilized to leverage parallel processing capabilities for both the training and testing phases.

In this paper, we investigate six CNN architectures: GoogLeNet, ResNet-50, Inception-v3, ResNet-101, DenseNet-201 and Our ESWCNN. Details about these architectures are provided in Table 2. Each model has been independently trained with the training data of the variant dataset.

**Table 2.** Summary the information about compared network architectures.

| Network | Depth | Number of Parameters(M) | Image Input size |
| --- | --- | --- | --- |
| GoogleNet [25] | 144 | 1.24 | $224 \times 224$ |
| SqueezeNet [23] | 68 | 1.23 | $227 \times 227$ |
| InceptionV3 [24] | 315 | 23.9 | $299 \times 299$ |
| ResNet-50 [26] | 50 | 25.6 | $224 \times 224$ |
| ResNet-101 [26] | 101 | 2.61 | $224 \times 224$ |
| DenseNet-201 [27] | 201 | 1.24 | $224 \times 224$ |
| **Our Model** | 7 | 0.06 | $28 \times 28$ |

The optimal hyperparameter values used in this study are presented in Table 3. Through experimentation, we determined that a batch size of 10 and a learning rate of 0.0003 complemented each other well in achieving our primary training objective of minimizing the generalization gap between training loss and validation loss.

**Table 3.** Hyperparameters in the Resnet50,Resnet101 and Densenet201 architectures.

| Hyperparameters | Values |
| --- | --- |
| Optimizer | Sgdm |
| Learning Rate | 0.0003 |
| Loss Function | Binary Cross-entropy |
| MiniBatchSize | 10 |
| MaxEpochs | 6 |
| Dropout | 0.2 |
| Shuffle | Every-epoch |
| Input size | $224 \times 224 \times 3$ |

Furthermore, a dropout rate of 0.2 was employed to prevent overfitting during model training. Subsequently, we saved the weights of the model with the lower validation loss. These saved weights were then utilized for ensembling and classifying the test images. It is noteworthy that we retained the default parameters for convolutional filters, padding, pooling filters, and strides from the original ResNet-50 and DenseNet-201 networks.

Structure from Motion (SfM) offers the capability to extract 3D surfaces, perform triangulation, and extract 3D features. Additionally, it enables the extraction of individual frames from videos. However, SfM typically requires videos in AVI format, while UCI dataset only provides videos in MP4 format. To address this discrepancy, a commercial software tool like Universal Format Factory (reference [49]) can be utilized for video conversion to ensure compatibility with SfM’s requirements.

On the GDZY datasets, experiments are conducted under uniform spatial and environmental conditions to compare the electromagnetic signals of the acquisition device’s signal line in normal operating and non-operating (power off) states. During the experiments, operators analyze the 1-10Hz frequency-domain and time-domain signals to detect any shooting signals being transmitted by the equipment and to identify weak magnetic signals from various patients.

Each test is performed three times in both normal working and non-working states (power off) within each test room, with measurements taken every minute. This results in a total of 24 tests conducted on a single individual, with each test lasting for 1 minute.

### Performance metrics

In the context of classification models, the recall rate is used as the evaluation metric. When dealing with colorectal polyp classification, which is a class imbalance problem, the performance of both individual and ensemble models is assessed using the F1-score metric. The F1-score provides equal importance to both precision and recall, making it an ideal metric for unbiased evaluation of performance in imbalanced datasets.

Given the dataset’s varying degrees of class imbalance, evaluating imbalanced data results necessitates the use of advanced metrics. Additionally, since the work involves three-class classification, the evaluation metrics include accuracy, precision, recall, specificity, precision, and F1-score.

In this study, the classification performance is evaluated based on sensitivity(sen), specificity(spe), and accuracy(acc). The terms True Positives (TP), False Positives (FP), True Negatives (TN), and False Negatives (FN) are defined as follows:

- True Positive (TP): A correctly classified image is considered a TP.
- False Positive (FP): An image that is not correctly classified is considered FP.
- True Negative (TN): The classifier estimates that the image class is not X, but actually represents the number of evaluations TN, which is not the image class X.
- False Negative (FN): The classifier estimated the image class not X, in fact the image class represents the number of evaluations in the form of X.

The following metrics are calculated from TP, FP, TN, FN:

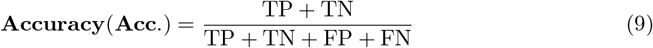

Specificity: the proportion of true negatives that were predicted as such. The specificity is given by:

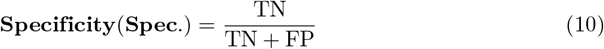

Sensitivity (or recall): the proportion of true positives that were predicted as such. The sensitivity is given by:

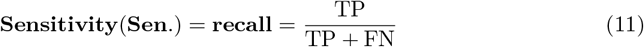

Precision (or PPV): the proportion of predicted positives that are real positives. The positive predictive value is given by:

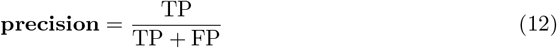

F1-score: a measure combining recall and precision. The F1-score is given by:

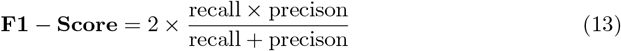

### Experiment Results and analysis

In the paper, the experimental settings and variations are presented in three tables: Table 4, Table 5, and Table 6. The evaluation metrics used to measure the performance of the models include accuracy, sensitivity, specificity, and F1-score.

**Table 4.** Ablation experiments on PolyGen.

| CNN | DWT | Sen. | Spec. | Acc. |
| --- | --- | --- | --- | --- |
| ✓ | × | 0.9117 | 0.9151 | 0.9317 |
| ✓ | ✓ | 0.9670 | 0.9283 | 0.94839 |

**Table 5.** Ablation experiments on UCI dataset.

| CNN | DWT | LSTM | Acc Hyp. | Acc Ade. | Acc Ser. | Sen Hyp. | Sen Ade. | Sen Ser. | Spec Hyp. | Spec Ade. | Spec Ser. |
| --- | --- | --- | --- | --- | --- | --- | --- | --- | --- | --- | --- |
| ✓ | × | × | **0.9871** | 0.9207 | **0.9790** | 0.9174 | **0.9975** | 0.9475 | 0.9725 | 0.9325 | **0.9575** |
| ✓ | ✓ | × | 0.9855 | **0.9511** | 0.9619 | **0.9250** | 0.9800 | **0.9875** | **0.9837** | **0.9563** | 0.9525 |
| × | ✓ | ✓ | 0.8250 | 0.7750 | 0.9750 | 0.8684 | 0.9688 | 0.7800 | 0.8750 | 0.8182 | 0.8000 |

**Table 6.** Ablation experiments on GDZY dataset.

| LSTM | DWT | PCA | FFT | Acc |
| --- | --- | --- | --- | --- |
| ✓ | × | × | × | 0.5583 |
| ✓ | ✓ | × | × | 0.5608 |
| ✓ | ✓ | ✓ | × | 0.6147 |
| ✓ | ✓ | ✓ | ✓ | **0.6741** |

These metrics are essential for assessing the classification models’ performance in the context of colorectal polyp classification, especially in dealing with class imbalance and multi-class classification scenarios. The tables provide a detailed overview of the experimental setups and variations tested in the study, along with the corresponding results based on the evaluation metrics mentioned.

### Ablation experiment

The experiments described in the paper were conducted using benchmark data and involved six contemporary CNN architectures. The main objective of these experiments was to identify the most suitable hyperparameter settings among the neural networks for classification tasks.

Initially, the dataset was divided into a training set and a test set. The training set was then fed into pre-trained networks to determine the optimal optimizer for the specific dataset. Three candidate optimizers were considered: ADAM (Adaptive Moment Estimation), SGDM (Stochastic Gradient Descent), and RMSprop (Root Mean Square Propagation). Through experimental comparison, it was found that SGDM performed the best among the three optimizers for the given dataset.

The process of evaluating different optimizers is crucial in determining the most effective optimization algorithm for training neural networks. The choice of optimizer can significantly impact the training process and ultimately influence the classification performance of the models. By comparing and selecting the best optimizer for a specific dataset and neural network architecture, researchers and practitioners can enhance the efficiency and effectiveness of the training process, leading to improved classification accuracy and overall model performance.

In the experiment, we conducted ablation experiments on the PolyGen dataset using a Convolutional Neural Network (CNN). The results of the experiments demonstrated that integrating Discrete Wavelet Transform (DWT) with CNN led to improvements over the original CNN model. Specifically, the CNN + DWT model exhibited enhancements in sensitivity, specificity, and accuracy by 5.6%, 1.3%, and 1.7% respectively, achieving values of 96.7%, 92.83%, and 94.8% for these metrics.

The ablation experiments involving CNN and DWT on the PolyGen dataset are summarized in Table 4. These findings highlight the effectiveness of combining CNN with DWT for classification tasks, showcasing significant performance gains in sensitivity, specificity, and accuracy compared to using CNN alone. The results underscore the potential advantages of leveraging both CNN and DWT techniques in tandem to enhance classification outcomes. CNN, DWT and LSTM combinations were used in the UCI database. The experimental results demonstrated that the combination of CNN + DWT was better than that of LSTM + DWT and the CNN model, but in some indicators, the model of Hyperplastic serrated accuracy, adenoma sensitivity, serated specificity and CNN was slightly ahead by 1 %. The combination of CNN and DWT performs better overall, with advantages in adenoma accuracy, hyperplastic and serrated sensitivity, and hyperplastic and adenoma specificity. In particular, it outperforms the CNN model by 3.1% in adenoma accuracy and by 4% in serrated sensitivity. LSTM excels in handling sequential information but does not have a comparative advantage in the various metrics compared.

Using CNN, DWT, and LSTM combinations for experiments on the UCI database, the results show that the CNN+DWT combination performs better overall than the LSTM+DWT combination and the standalone CNN model. However, in some individual metrics such as Hyperplastic serrated accuracy, adenoma sensitivity, and serrated specificity, the CNN model slightly outperforms by 1%. The CNN+DWT combination excels in adenoma accuracy, hyperplastic and serrated sensitivity, and hyperplastic and adenoma specificity, with notable leads in adenoma accuracy and serrated sensitivity, surpassing the CNN model by 3.1% and 4% respectively. Although LSTM is better at handling sequential information, it does not demonstrate superiority in the comparison of various metrics.

Ablation experiments (DWT,CNN,LSTM)on PolyGen shown in Table 5 On the GDZY database, a combined experiment was conducted using LSTM, DWT, PCA, and FFT. Since the sampled data consisted of weak magnetic sequence signals, LSTM was used instead of CNN. The accuracy of LSTM alone was 55.83%. With the addition of DWT and PCA, the accuracy increased to 56.8% and 61.4% respectively. The final combination achieved an accuracy of 67.4%. Ablation experiments (DWT,PCA,FFT,LSTM)on GDZY shown in Table 6

In Equation 8, the parameter *γ* is used to balance CNN and scattering wavelet. An appropriate *γ* value was determined through experiments. In the grouped experiments, we set *γ* = 0, 0.25, 0.5, 0.75, 1, the variant *γ* value from Eq. 8 are used in experiment shown in Fig 10, The experiments compared the average accuracy, hyperplastic accuracy, adenoma accuracy, and serrated accuracy, and the results showed that *γ* = 0.5 achieved the best experimental performance. In subsequent experiments, we used this *γ* = 0.5 value as the experimental setting.

**Fig 10.**
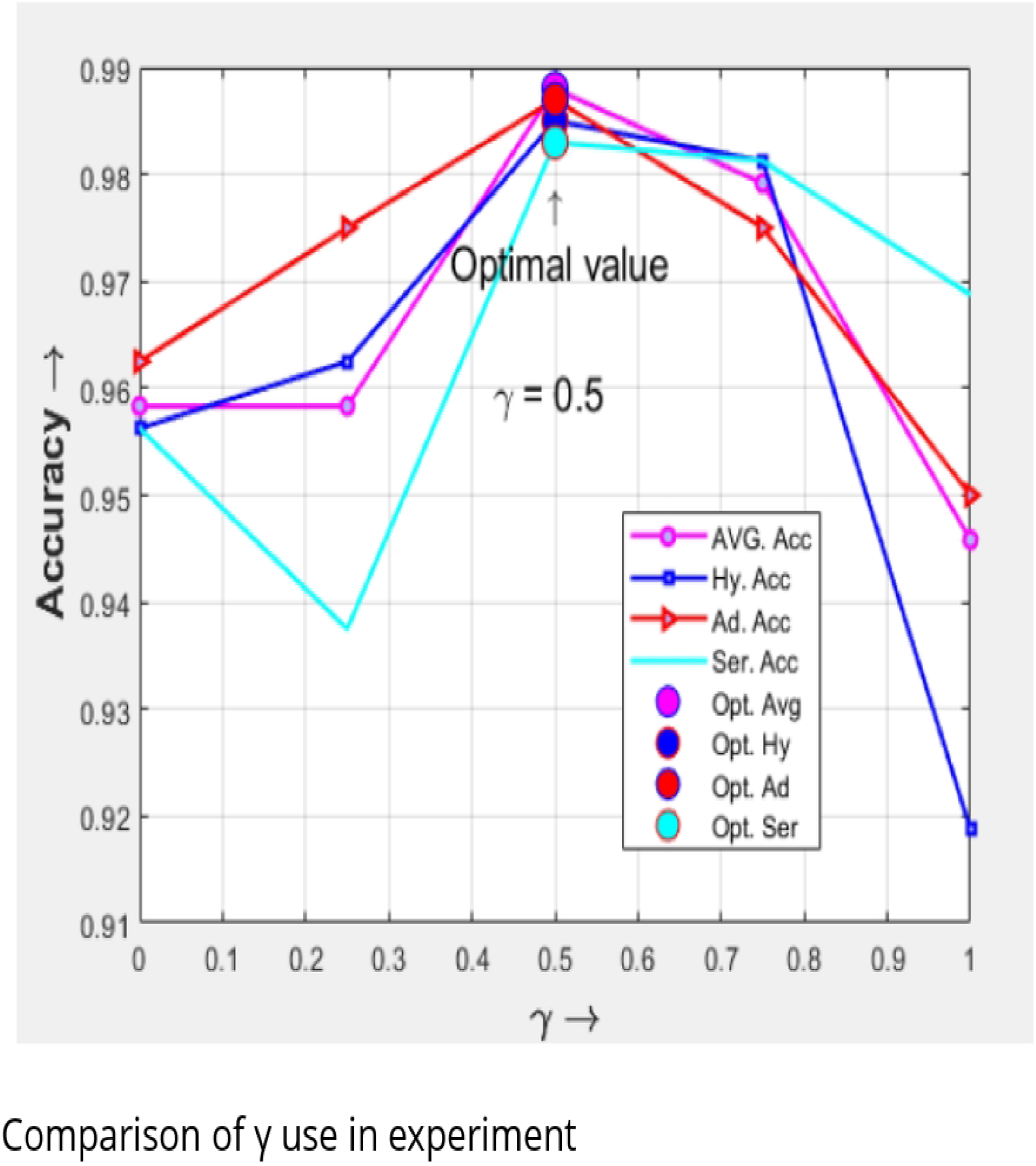
Comparis on of, use in experim ent.

### Frame-based three-class polyp classification

In the UCI dataset, the methods were classified into three classes: adenoma, hyperplastic, and serrated. Another commonly used technique in the literature to enhance performance is feature selection or reduction algorithms. In this study, ESWCNN was proposed as a feature reduction method. Table 7 presents the classification performance of different feature selection algorithms in the CNN classifier and time measurements for a feature vector. As shown in Table 7, ESWCNN improved the classification performance of CNN architectures from 95.4% to 96.4%. Additionally, the implementation time of the Discrete Wavelet Transform (DWT) architecture was found to be shorter compared to other methods. Table 8 illustrates the computational complexity of the proposed method.

**Table 7.** Comparison of three-class polyp classification on UCI dataset.

| Algorithm | Average<br>Acc.(%) | Acc<br>Hyp.(%) | Acc<br>Ade.(%) | Acc<br>Ser.(%) |
| --- | --- | --- | --- | --- |
| CAC [43] | 82.4 | 89 | 76 | 87 |
| Deep feature selection [51] | 94.8 | 98 | 95 | 89 |
| Genetic [50] | 94.7 | 98 | 96 | 89 |
| Differential evolution [50] | 94.6 | 97 | 95 | 89 |
| PCA [50] | 93.6 | 97 | 93 | 89 |
| LDA [50] | 94.7 | 98 | 96 | 89 |
| PCC [52] | 94.4 | 98 | 95 | 89 |
| F-score [52] | 94.6 | 98 | 95 | 89 |
| CNN+DWT+SVM [33] | 95.4 | 98 | 96 | 90 |
| ESWCNN | **96.4** | **98.5** | **96.2** | **95.1** |

**Table 8.** classification accuracy with others CNNs algorithms.

| Model | Average<br>Acc.(%) | Acc<br>Hyp.(%) | Acc<br>Ade.(%) | Acc<br>Ser.(%) | Times.(s) |
| --- | --- | --- | --- | --- | --- |
| Squeezenet [23] | 33.04 ± 3.56 | 100 | 0.00 | 0.00 | **314.04 ± 13.4** |
| Googlenet [25] | 58.75 ± 3.56 | 72.50 ± 4.86 | 28.75 ± 3.34 | 75.01 ± 2.23 | 592.19 ± 54.3 |
| InceptionV3 [24] | 90.0 ± 9.21 | 87.50 ± 4.86 | 95.0 ± 3.34 | 87.50 ± 2.54 | 5217.5 ± 67.7 |
| Resnet 50 [26] | 69.06 ± 3.86 | 82.50 ± 3.63 | 18.75 ± 4.12 | 76.25 ± 6.73 | 1653 ± 173.1 |
| Resnet 101 [26] | 73.12 ± 7.10 | 85.00 ± 3.46 | 33.75 ± 2.91 | 92.50 ± 8.46 | 2896 ± 397.8 |
| Densenet 201 [27] | 96.25 ± 1.42 | 96.25 ± 4.27 | **97.5 ± 12.5** | 95.0 ± 3.68 | 3748 ± 335.3 |
| ESWCNN | **96.4 ± 4.36** | **98.51 ± 2.25** | 96.20 ± 12.5 | **95.1 ± 2.57** | 811 ± 72.1 |

When compared with other CNN architectures, Fig 11 graphically represents the mean values for all metrics, including accuracy, precision, recall, specificity, and F1 score. The comparison involves ESWCNN, which combines ResNet101 and ResNet50 architectures, with the DenseNet201 architecture.

**Fig 11.**
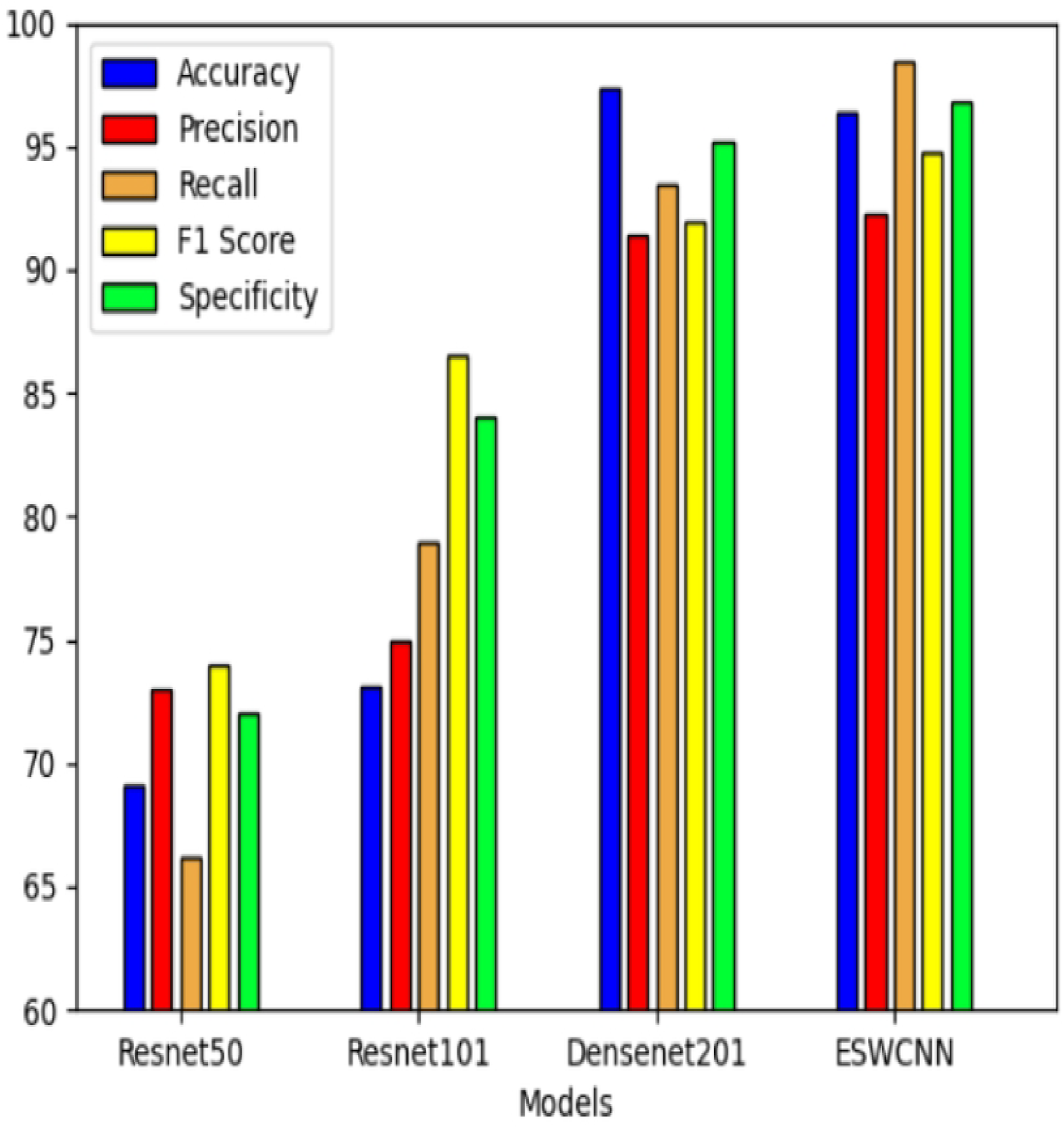
Test results of all four classifiers for pol_yp_s classification

As shown in Table 8, the proposed model demonstrated a minimum 55% improvement in processing time. Additionally, ESWCNN was observed to enhance the average accuracy rate. Conversely, DWT exhibited faster processing speeds compared to all other CNNs, attributed to its feature size reduction capabilities, consequently boosting the classifier mean accuracy. The comparison of classification accuracy with other CNN algorithms is presented in Table 8.

In terms of overall performance, Densenet stands out by achieving the highest accuracy in adenoma classification at 97.5%, surpassing ESWCNN’s 96.2%. Although other metrics are comparable to ESWCNN, the depth and larger input image size of Densenet result in a significantly longer average time required for 5-fold cross-validation on the UCI dataset, reaching 3748 seconds, which far exceeds ESWCNN’s 811 seconds. On the other hand, models like Resnet 50 exhibit an average time of 1653 seconds with an accuracy of only 69%. Googlenet and Squeezenet, while demonstrating lower average processing times than ESWCNN, have accuracies of only 58% and 33%, respectively.

### Frame-based two-class polyp classification

From Table 9, it is evident that the average accuracy, sensitivity, specificity, and run times for the 5-fold cross-validation experiment on the PolyGen dataset are superior. This indicates that the models excel in correctly predicting polyp categories.

**Table 9.**
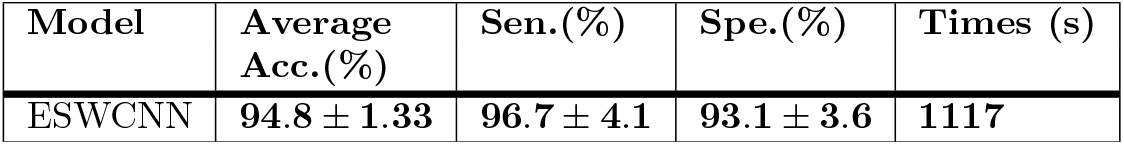
n-fold cross validation Experiment result on PolyGen.

In the classification experiment conducted on PolyGen, a total of 6000 images were resized to 28 *×* 28 pixels. The image classification models proposed in this study utilized a 5-fold cross-validation method to split the dataset into training (80%) and test data (20%). The experiment yielded an average accuracy of 94.8% with a standard deviation of 1.33. The sensitivity was measured at 96.7%, while the specificity was at 93.1%.

The results presented in Table 9 demonstrate the performance achieved when concatenating 10 scattering wavelet layers with 10 CNN layers.

The experiment results for classification using different CNNs are summarized in Table 10. Densenet 201 stands out with the highest accuracy of 95.83% among all CNN models, which aligns with the results obtained in the two-class classification scenario, result shown in Fig 12.

**Table 10.** Experiment result with CNNs on PolyGen.

| Model | Data size | Accuracy (%) | # of Iteration | Times (s) |
| --- | --- | --- | --- | --- |
| Restnet 50 | Original image,not imresize | 86.77 | 1440 | 8125 |
| Restnet 101 | Original image,not imresize | 78.39 | 2880 | 14,764 |
| Densetnet 201 | Original image,not imresize | 80.46 | 2880 | 17,219 |
| Restnet 50 | imresize to $224 \times 224$ | 93.50 | 1440 | 7619 |
| Restnet 101 | imresize to $224 \times 224$ | 84.92 | 2880 | 14,179 |
| Densetnet 201 | imresize to $224 \times 224$ | **95.83** | 2880 | 16,163 |
| ESWCNN | imresize to $28 \times 28$ | 94.83 | **1000** | **1117** |

**Fig 12.**
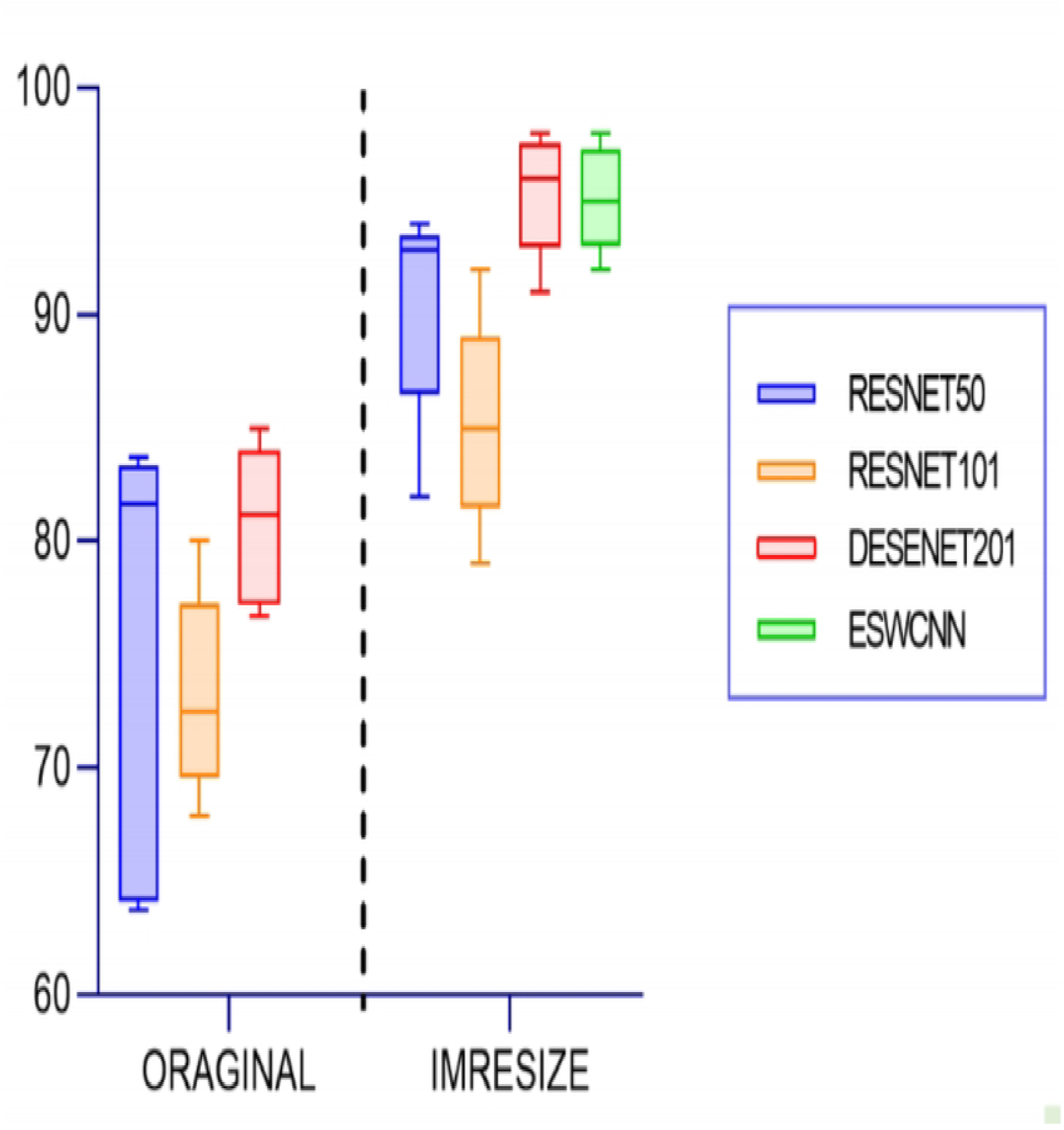
Polyp classification comparison result on PolyGen

**Fig 13.**
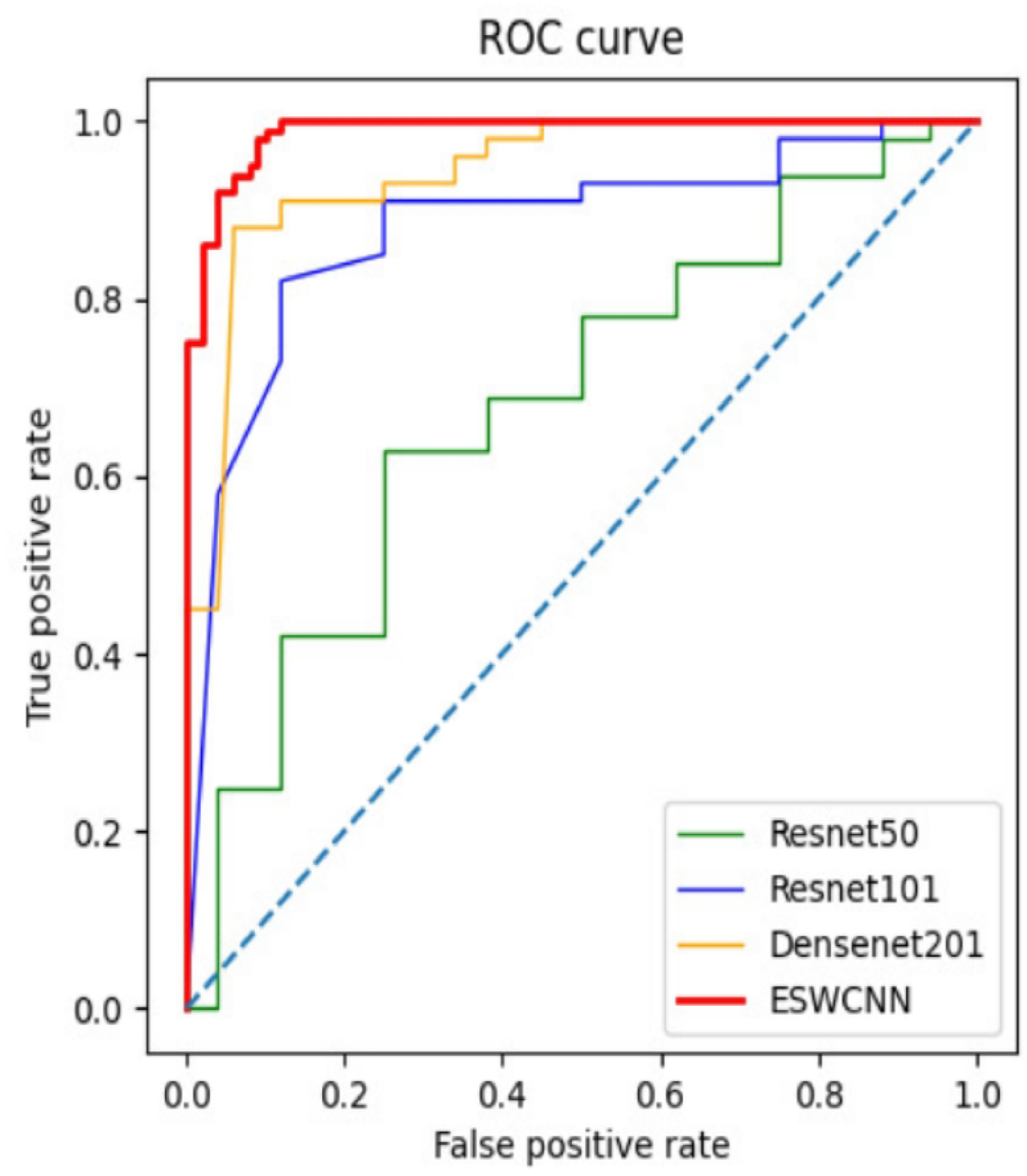
Area under the ROC curve analysis for pol_yp_ classification

Comparatively, Densenet boasts a deeper network architecture with 201 layers. On the other hand, ESWCNN achieved a classification accuracy of 94.83% with a relatively shallower network structure, showcasing a marginal difference of less than 1% compared to Densenet. Interestingly, ESWCNN also demonstrated a significantly lower computation time of 1117 seconds, in stark contrast to the 16163 seconds required by Densenet.

Under the Resnet configuration, the accuracy achieved was 93.5%, with a computation time of 7619 seconds. Notably, ESWCNN exhibited enhanced computational efficiency compared to Resnet.

In the GDZY dataset, a total of 1347 samples were provided. Some samples with incomplete data were removed, resulting in the selection of 1180 samples for analysis. Among these, 944 samples were allocated for training purposes, while the remaining 236 samples were reserved for testing. The experimental setup involved utilizing a 5-fold cross-validation technique. The experimental outcomes for ESWCNN on the GDZY dataset are as follows:

- Accuracy: 77.5%
- Sensitivity: 80%
- Specificity: 75.6%

ESWCNN was compared against FFT+PCA and XGBoost methodologies. The experiment results shown in Table 11, including the confusion matrices, for the experiments conducted on the GDZY dataset were analyzed.

**Table 11.** Confusion matrices for Experiment result on GDZY.

|  | ESWCNN |  | XGBoost |  | FFT+PCA |  |
| --- | --- | --- | --- | --- | --- | --- |
|  | Positive | Negative | Positive | Negative | Positive | Negative |
| Positive | 81 | 33 | 63 | 34 | 55 | 37 |
| Negative | 20 | 102 | 38 | 101 | 46 | 98 |
| Sum | 101 | 135 | 101 | 135 | 101 | 135 |

In the grid search process conducted to optimize the XGBoost model’s hyperparameters, the following ranges and options were explored to identify the best-performing configuration in terms of accuracy, recall, and specificity:

- Max depth: Ranging from 3 to 11
- Learning rate: Options included 0.1, 0.01, and 0.001
- Number of estimators: Choices were 10, 30, 70, 150, 250, and 500
- Sampling ratios: Ranging from 0.5 to 1
- Subsampling ratios: Varied from 0.5 to 1

The grid search process aimed to identify the combination of these hyperparameters that led to the highest performance metrics, such as accuracy, recall, and specificity, for the XGBoost model when applied to the GDZY dataset.

## Discussion

In the context of UCI classification, our proposed ESWCNN achieved average accuracies of 96.4%, 98.5%, 96.2%, and 95.1% for overall accuracy, hyperplastic accuracy, adenoma accuracy, and serrated accuracy, respectively. Additionally, for the PolyGen 2-class poly classification, the ESWCNN model attained average accuracies of 94.8%, sensitivity of 96.7%, and specificity of 93.1%.

Furthermore, a receiver operating characteristic (ROC) analysis was conducted to evaluate the model performance, and Figure 9 illustrates the results using the area under the ROC curve (AUC). It is clear from this analysis demonstrated that our proposed ESWCNN outperforms all other convolutional neural networks (CNNs) for both classification tasks, showcasing its superior performance and effectiveness in handling the given classification problems.

In our study, we emphasize the importance of minimizing both false positives (FP) and false negatives (FN) due to their critical impact on the accuracy of the classification system. False negatives occur when patients with cancerous tumors are incorrectly labeled as noncancerous, while false positives occur when patients without cancerous tumors are inaccurately classified as abnormal (cancerous). Both FP and FN can lead to misdiagnosis, posing significant risks to human health.

To address this issue, we have incorporated the F1 score along with other performance evaluation metrics to give equal importance to both FP and FN. By considering a balanced evaluation approach, we aim to reduce the occurrence of misclassifications and enhance the overall diagnostic accuracy of our proposed ESWCNN model.

Furthermore, we have provided visual representations of the lesions that were correctly classified by both human experts and our ESWCNN model, as well as those that were misclassified by the model but correctly identified by the human experts. Additionally, the images of wrongly classified polys are also presented in Fig 14, offering a comprehensive overview of the classification outcomes for further analysis and discussion.

**Fig 14.**
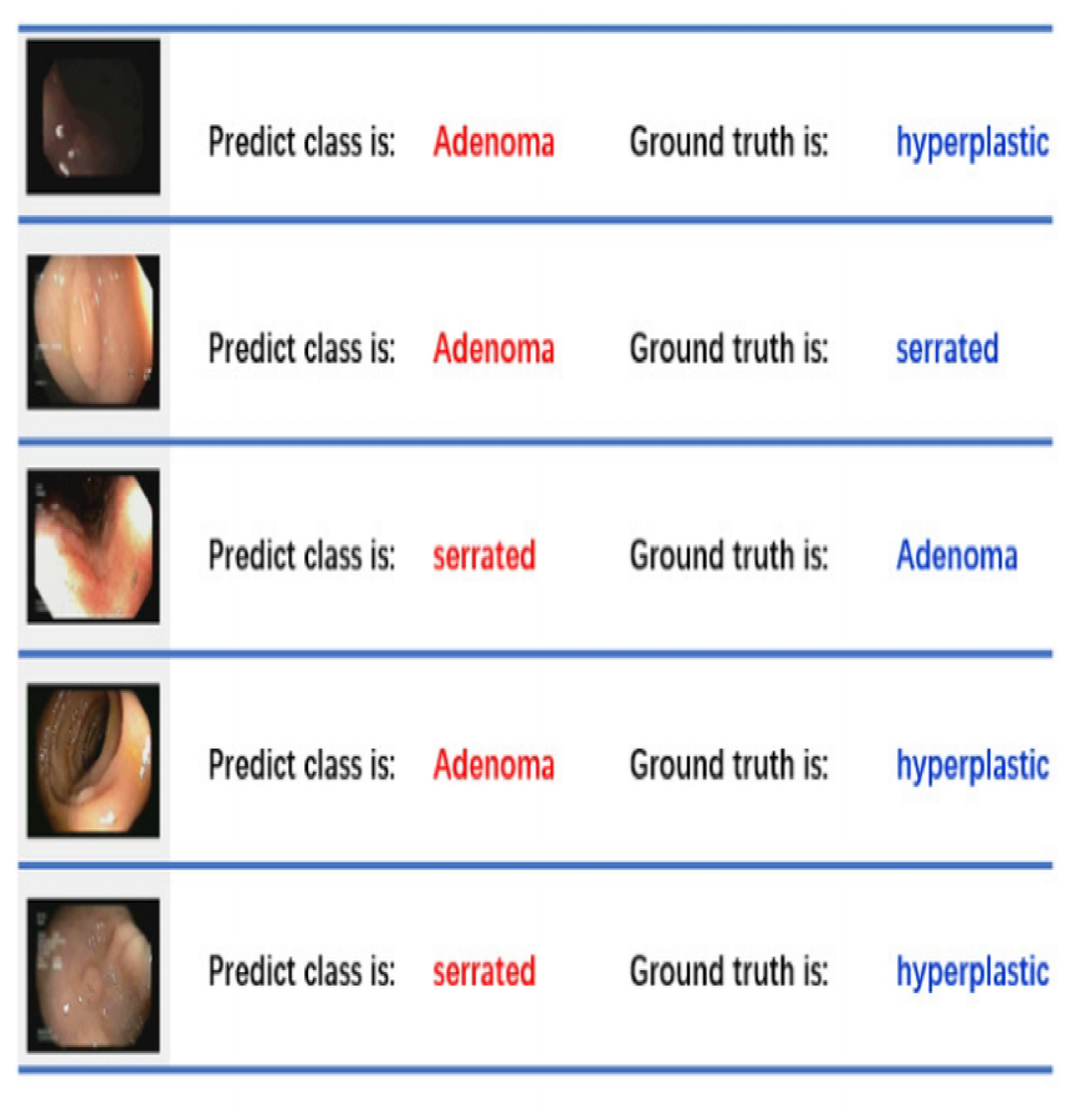
Some sample wrongly classified on UCI dataset.

In a few instances of misclassified samples, we observed that the ground truth label “hyperplastic” is susceptible to being incorrectly classified as “adenoma.” Similarly, there are mutual misclassifications between “adenoma” and “serrated” labels. These misclassifications indicate potential challenges in accurately distinguishing between these different types of lesions, highlighting the complexity and nuances involved in the classification task. Further investigation and refinement of the classification model may be necessary to address these specific misclassification patterns and improve the overall accuracy of the system.

The samples in the UCI database are categorized as positive or negative classifications by the ESWCNN model. The best model, as illustrated in Fig 15, accurately classifies the lesions in the dataset.

**Fig 15.**
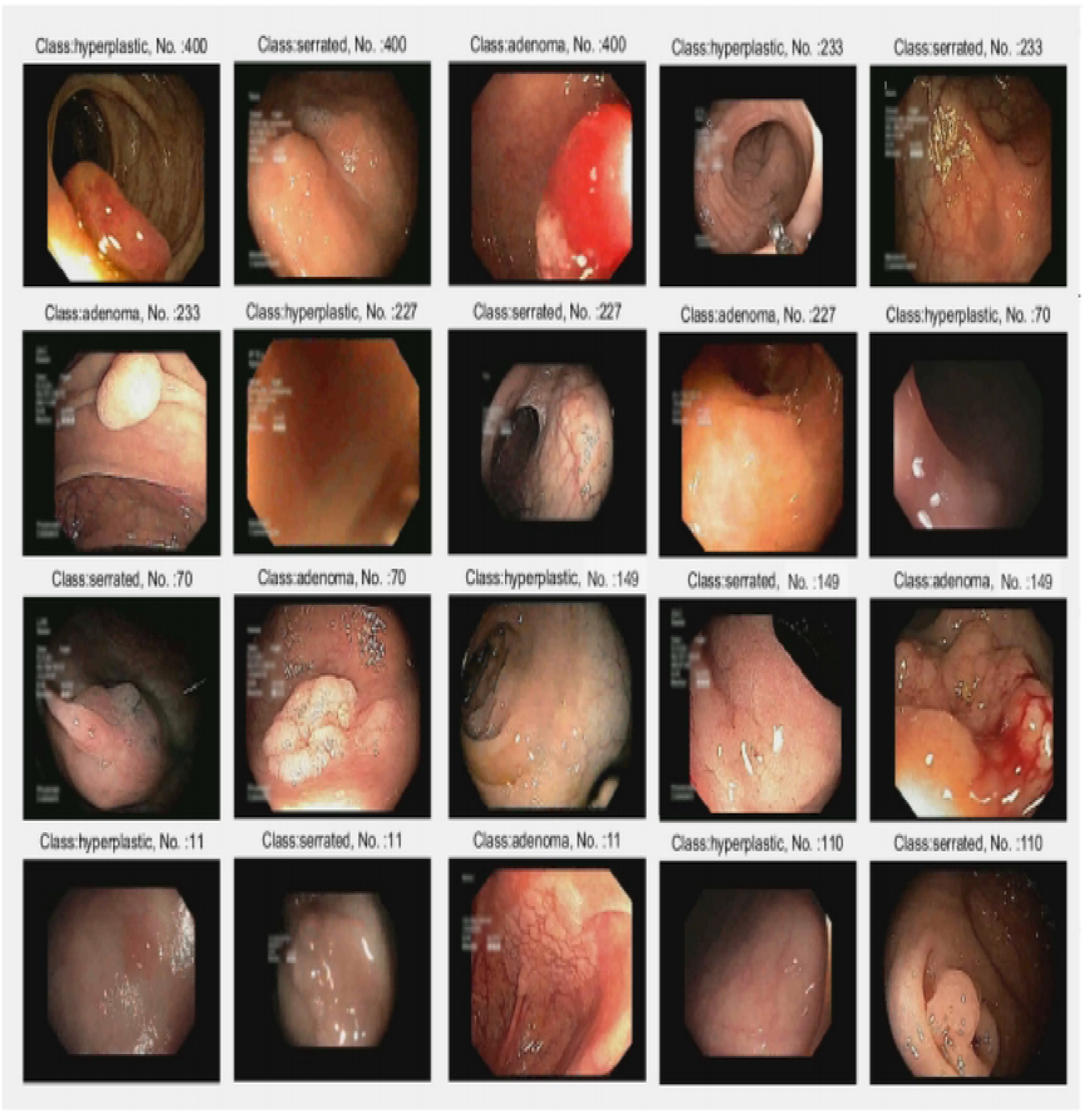
Some sample correctly classified on UCI dataset.

## Conclusion

There are several significant challenges in medical image processing when utilizing CNN models. The high computational cost arises from pixel-level operations, making it a critical issue. Deep learning algorithms typically involve millions of parameter updates during training, necessitating expensive hardware resources such as high-end graphics processing units. Moreover, obtaining labeled data for medical images is a challenging task, as it requires substantial time from medical professionals and multiple expert opinions to minimize human error. These obstacles hinder the application of high-performance algorithms like deep learning in polyp classification.

To address these challenges, we propose ESWCNN model which combines simple CNN architectures with scattering wavelets to extract features from polyp images. ESWCNN leverages CNN for spatial feature extraction and scattering wavelets for frequency feature extraction, updating parameters through backpropagation. We conducted experiments on two public databases and one private database, including the UCI database with three colorectal polyp categories, and the PolyGen and GDZY databases with two categories each. Our experiments involved ablation studies, comparisons with state-of-the-art (SOTA) methods, and evaluations against commonly used CNN architectures. Various parameter configurations were tested, resulting in significant improvements across all experimental metrics.

On the UCI database, the accuracy improved from 95.4% to 96.4%. ESWCNN outperformed traditional CNN models in classifying Hyperplastic, Adenoma, and Serrated polyps with accuracies of 98.5%, 96.2%, and 95.1% respectively, while requiring only 25% of the time compared to Densenet. On the PolyGen database, ESWCNN achieved superior performance compared to ResNet and Densenet, with an accuracy of 94.83% and a completion time of 1117 seconds. On the GDZY database, the results were as follows: Accuracy: 77.5%, Sensitivity: 80%, Specificity: 75.6%.

The experimental outcomes demonstrate the efficiency and performance advantages of our proposed method in colorectal polyp classification, surpassing SOTA methods. This suggests potential value for future clinical applications.

## Data Availability

1,UCI public dataset: Mesejo, P., Pizarro, D., Abergel, A., Rouquette, O., Beorchia, S., Poincloux, L., Bartoli, A. (2016). Computer-aided classification of gastrointestinal lesions in regular colonoscopy. IEEE Transactions on Medical Imaging. http://www.depeca.uah.es/colonoscopy_dataset/ 2, PloyGen data set Ali S, Dmitrieva M, Ghatwary N, Bano S, Polat G, Temizel A, et al. Deep learning for detection and segmentation of artefact and disease instances in gastrointestinal endoscopy. Medical Image Analysis. 2021:102002. https://www.synapse.org/#!Synapse:syn45200214 3,GDZY Data cannot be shared publicly because of Ethics. Data are available from the the Second Affiliated Hospital of Guangzhou University of Traditional Chinese Medicine(TCM) Institutional Data Access / Ethics Committee.

## Acknowledgments

This work was supported by Guangdong Province Key Laboratory of Computational Science at the Sun Yat-sen University (2020B1212060032), and the National Science Foundation of China (11971491, 12171490,62376291), and the Foundation of Guangdong-Hong Kong-Macao National Center for Applied Mathematics (2021B1515310002).

## Notes

### Competing Interest Statement

The authors have declared no competing interest.

### Funding Statement

Yes

### Author Declarations

We have obtained ethical committee approval, from Ethics Committee of Guangdong Provincial Hospital of Chinese Medicine，Approval Letter No.: Ethics Committee of Guangdong Provincial Hospital of Traditional Chinese Medicine BE2020-082-01, Review date: First review: April 30,2020 Review: June 15,2020 All patient data has been analyzed anonymously, and no patient names or other private information will appear in the paper.

